# Brick kilns, child respiratory symptoms and blood lead in Bangladesh: a national geospatial analysis of the 2025 Multiple Indicator Cluster Survey

**DOI:** 10.64898/2026.10.22.26366042

**Authors:** S. M. Ashraf Ul Hoque, Mohammad Khalid Hasan, Md Khairul Islam, Md. Mohiuddin Sharif, Suraiya Akter Jemy, S M Raysul Haque, Kamran ul Baset

**Author notes:** Corresponding author: S. M. Ashraf Ul Hoque, Department of Public Health, Independent University Bangladesh (IUB), Plot 16 Aftab Uddin Ahmed Rd, Bashundhara Residential Area, Dhaka 1229, Bangladesh.

## Abstract

**Background:** Brick kilns are a major source of air pollution in Bangladesh. Their effects on child respiratory health and lead exposure have not been studied nationally with blood measures.

**Methods:** We linked a 2018 to 2019 satellite map of 6,978 kilns to the 3,149 displaced clusters of the Bangladesh Multiple Indicator Cluster Survey 2025. Exposure was the number of kilns within 5 km. Outcomes were cough and acute respiratory infection (ARI) symptoms in 23,357 children under 5 years, and venous blood lead in 10,622 children aged 12 to 59 months. We fitted survey-weighted modified Poisson models with district fixed effects. Diarrhoea and interview timing relative to the firing season served as negative controls.

**Findings:** Three in four children (75.4%, 95% CI 73.6 to 77.1) lived within 5 km of a kiln, and 20.3% (18.7 to 22.0) had ten or more. Exposure rose with wealth. Compared with no kiln, ten or more kilns were not associated with cough (adjusted prevalence ratio [PR] 1.05, 0.96 to 1.16), ARI symptoms (1.21, 0.90 to 1.62) or blood lead of 5 µg/dL or more (0.94, 0.85 to 1.03; crude PR 1.27, 1.14 to 1.41). Per 10 kilns, the ARI PR was 1.11 (1.01 to 1.21), but a spline showed no association (P=0.153). Cough was not associated with kiln density at the peak of firing (PR per 10 kilns 1.00, 0.96 to 1.05) but was in May and June (1.10, 1.04 to 1.17). The lead result held in all sensitivity analyses.

**Interpretation:** At survey resolution, living among more kilns was not linked to blood lead. Weak respiratory associations, absent at peak firing, fit confounding by place better than an acute effect of kiln smoke. Studies with exact locations should measure effects within 1 to 2 km of kilns.

**Funding:** None.

## Introduction

Bangladesh produces bricks in about 7,000 kilns that burn coal and biomass during the dry season [1,2]. Kilns emit fine particles, black carbon, sulphur dioxide and metals [3,4]. Models attribute 30 to 50% of winter fine particles in Dhaka to kilns [1,3], and source apportionment in Dhaka has also identified kilns as a major source [5]. A chemical transport model estimated that kilns added 31 to 46 µg/m³, or 18 to 27%, of monthly fine particulate matter across Bangladesh in January 2019 [6]. Satellite mapping found that at least 12% of the population lives within 1 km of a kiln [1]. The Brick Manufacturing and Brick Kiln Establishment (Control) Act of 2013 bars kilns within 1 km of schools, health facilities and other protected sites [1]. Yet the same study found 77% of kilns within 1 km of a school.

Young children are the group most vulnerable to this pollution [7]. Pneumonia remains a leading cause of death in children under 5 years [8]. In Dhaka, childhood pneumonia rose with daily fine particles, more so on days when a larger share came from brick kilns [9]. Kilns may also add metals to the local environment [4]. Soil around kilns in Natore, a district in north-western Bangladesh, had lead above regional reference values, although that study did not attribute the excess to kiln emissions [10]. Blood lead is high in Bangladeshi children: 38.4% of children aged 12 to 59 months had 5 µg/dL or more in 2025 [11].

Evidence on kilns and child health is thin. A field study in Bangladesh linked downwind proximity during the production season to adult respiratory symptoms, but its estimates for young children were imprecise [2]. Small studies compared children living at kiln sites in Pakistan [12], or attending a school near kilns in Nepal [13], with other children. Our search of PubMed and Europe PMC found no study that used a national sample or measured blood lead alongside kiln exposure. The Bangladesh Multiple Indicator Cluster Survey (MICS) 2025 now offers cluster coordinates, respiratory symptoms and venous blood results for a national sample of young children [11]. Its fieldwork ran from February to June 2025, which spans the peak and the end of the firing season. This timing allows a check of whether associations follow the months in which kilns burn.

Our overall objective was to assess whether living near brick kilns is linked to respiratory symptoms and blood lead in young children in Bangladesh. We first estimated how many children live near kilns, by division, residence and wealth. We then examined cough and symptoms of acute respiratory infection, using diarrhoea and the timing of the firing season as checks for confounding. Finally, we examined blood lead in children aged 12 to 59 months, with household soil lead as a check on the deposition pathway.

## Methods

### Study design and data

This cross-sectional study linked the Bangladesh MICS 2025 [11] to a satellite map of brick kilns. The Bangladesh Bureau of Statistics conducted the survey with UNICEF support. Urban and rural areas of each district formed the sampling strata; 3,149 enumeration areas were selected with probability proportional to size, and 20 households were drawn in each. Fieldwork ran from 24 February to 30 June 2025. Teams of five female interviewers, a measurer, a phlebotomist and a supervisor collected data on tablets with the standard MICS questionnaires. One household per cluster was re-interviewed, and field check tables were reviewed weekly. Questionnaires were completed for 23,357 of 24,680 listed children under 5 years (94.6%). We followed the Strengthening the Reporting of Observational Studies in Epidemiology (STROBE) guideline [14].

In each household, one child aged 12 to 59 months was selected for venous blood collection. Of 17,132 eligible children, 10,661 gave blood (65.3%). The International Centre for Diarrhoeal Disease Research, Bangladesh (icddr,b) was the technical partner for collecting and testing blood and soil samples. Blood was tested in designated reference laboratories with validated methods, and a soil sample from the household environment of each child who gave blood was analysed with a portable X-ray fluorescence analyser [11]. The report does not name the blood assay or its detection limits.

### Kiln locations and exposure

The main kiln map came from Lee and colleagues, who applied deep learning to high-resolution satellite images from October 2018 to May 2019 [1]. It located 6,978 kilns, classified as 5,011 zigzag and 1,967 fixed-chimney kilns, with 94.2% accuracy [1] (figure 1A). Two lie more than 2 km outside the national boundary; we kept them in exposure counts but not on the map. The coordinates are public under a CC0 licence [15]. As a second map, we used 4,760 kilns from the Air Pollution Asset Data (APAD) Indo-Gangetic Plain kiln dataset, which maps kilns in six of the eight divisions of Bangladesh [16,17]. Its Bangladesh layer is released without removal of false positives [17].

**Figure 1.**
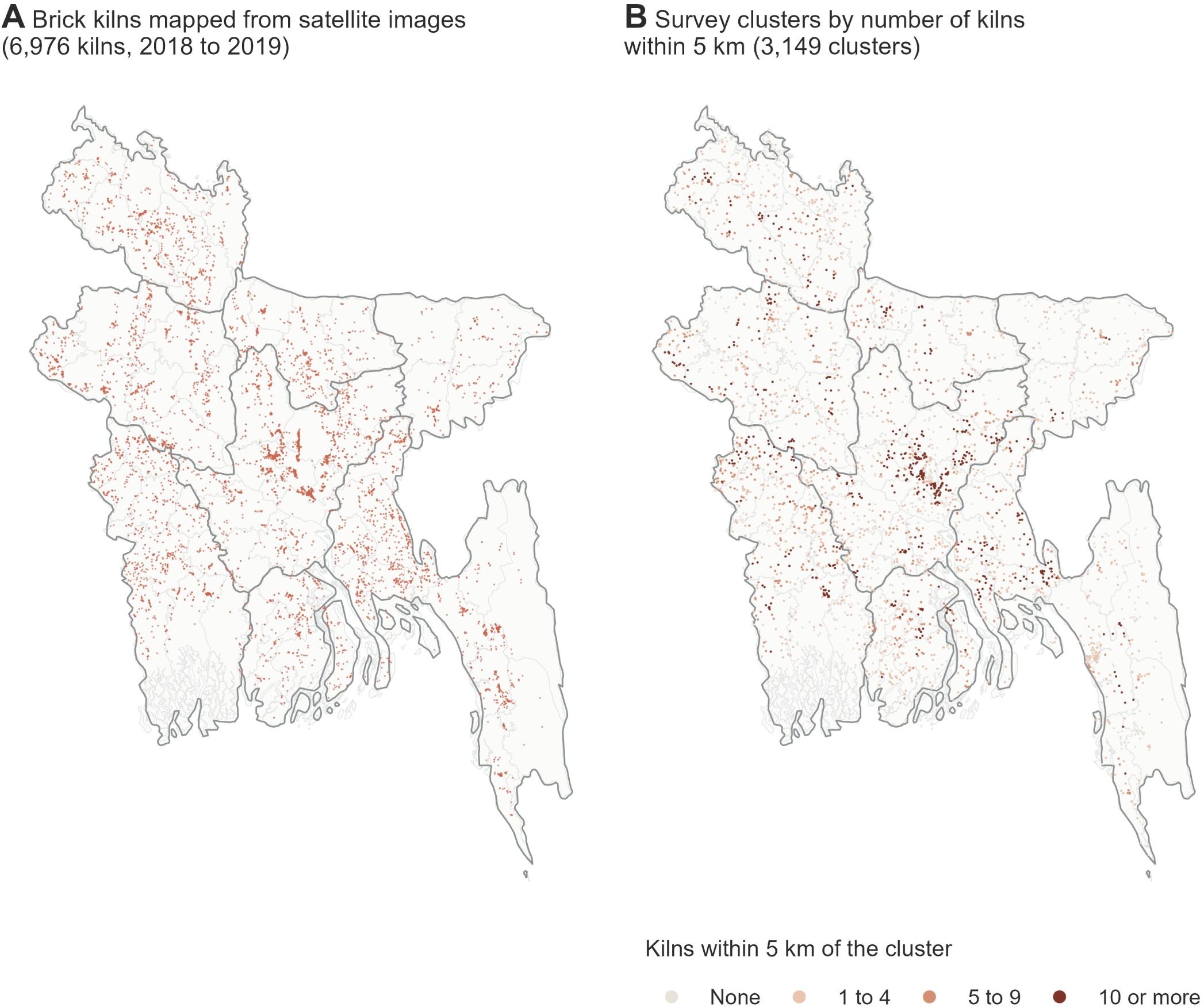
Brick kilns and survey clusters, Bangladesh. (A) Locations of 6,976 brick kilns inside Bangladesh mapped from satellite images taken between October 2018 and May 2019 [1]. (B) MICS 2025 survey clusters (displaced locations) coloured by the number of kilns within 5 km. Thin grey lines are district boundaries and dark grey lines division boundaries.

To protect respondents, MICS publishes cluster coordinates after random displacement within the upazila (subdistrict), with a procedure based on that of the Demographic and Health Surveys [18,19]. Urban clusters move up to 2 km and rural clusters up to 5 km, with 1% of rural clusters moved up to 10 km [19]. Distance analyses should therefore use buffers at least as wide as the displacement [19,20].

Our main exposure was the number of kilns within 5 km of the cluster point. We grouped it as none, 1 to 4, 5 to 9 and 10 or more (figure 1B). We also used the count per 10 kilns, the distance to the nearest kiln and the count within 10 km. Distances were computed in a projected coordinate system (Universal Transverse Mercator zone 46N). The main map covers Bangladesh only, so kilns across the border are missing for clusters near it. We therefore computed each cluster’s distance to the land border with India or Myanmar. We also counted consensus kilns: kilns in the main map with a kiln of the second map within 500 m.

### Outcomes

Mothers or caregivers reported whether the child had cough, difficult breathing and diarrhoea in the 2 weeks before the interview. Symptoms of acute respiratory infection (ARI) followed the MICS 2025 definition: fast or difficult breathing perceived to be due to a problem in the chest, or in both the chest and a blocked or runny nose. Unlike earlier rounds, this definition does not require cough [11]. Diarrhoea was the negative control outcome [21]. Household cooking smoke has been linked to childhood diarrhoea [22], and we adjusted for cooking fuel. A short-term effect of outdoor kiln smoke on diarrhoea is less plausible, so an association would point mainly to shared confounding.

For blood lead we used 5 µg/dL or more as the main cut-off, as in the survey report and WHO guidance [11,23]. We also analysed 3.5 µg/dL or more and log-transformed lead. Blood cadmium, arsenic and mercury were analysed as exploratory outcomes only. The survey report did not publish them, because detection thresholds, analytical precision and reference thresholds did not allow robust estimates [11]. Household soil lead (mg/kg) served as a marker of local deposition.

### Covariates

We chose confounders before analysis as factors that may influence both where families live relative to kilns and child health. These were child age group and sex, mother’s education, household wealth quintile, urban or rural residence, solid cooking fuel and month of interview. Kilns cluster just outside large urban areas [1], so we added log population density and log night-time lights from the MICS geospatial covariates [24]. District fixed effects compared clusters within the same district. For lead outcomes, a further model added lead sources reported in the blood module. These were smoking by a person in regular contact with the child, pica and lead-related activities near the home. They also included a household contact working with lead and use of sindoor (a vermilion powder) or surma (an eye cosmetic).

### Statistical analysis

All estimates accounted for survey weights, clustering and stratification with Taylor linearisation. Symptom analyses used child weights. Blood outcomes used the blood sample weights, which account for the selection of one child per household and for non-response within strata [11]. We first reproduced the national prevalence of diarrhoea, ARI symptoms and blood lead in the survey report.

For the first aim, we estimated the weighted share of children with at least 1 kiln and with 10 or more kilns within 5 km. We also estimated the share whose nearest kiln was under 2 km away. We reported these shares by division, residence and wealth quintile.

For the second and third aims, we estimated prevalence ratios (PR) with modified Poisson regression [25]. Log lead was modelled with linear regression. The primary outcomes were cough, ARI symptoms and blood lead of 5 µg/dL or more, and the primary exposure was the kiln category within 5 km. Other outcomes and exposure measures were secondary. We report crude and adjusted estimates and a test for trend that scores the categories 0 to 3. To show which adjustments changed the estimates, we added covariates in steps: child and household factors, then urban setting and population measures, then division and finally district. The analyses produced many estimates, so we read isolated results with P values near 0.05 as hypothesis-generating.

We used the season of firing as a negative control period. Kilns fire in the dry months, from about November to April or May [1,26]. We grouped interviews into February to March (peak firing), April (late firing) and May to June (end of firing and after). A true effect of kiln smoke on recent symptoms should be present during firing and weaken after it. We fitted kiln-by-period interactions, with kilns as categories and per 10 kilns, and report period-specific estimates.

We planned three sensitivity analyses. First, we used other exposure measures: nearest-kiln distance and kiln counts within 10 km. Second, we repeated the analysis with the second kiln map, comparing both maps within the six divisions it covers. Third, we restricted the analysis to urban clusters, which are displaced by at most 2 km. To judge how displacement affects exposure, we displaced each published point a second time with the survey rule, 100 times, and recomputed exposure, overall and within districts.

After review of a first draft, we added post hoc analyses (S1 Table). We fitted a restricted cubic spline of the 5 km count and top-coded the count at its 99th centile. We excluded clusters within 5 km of the land border and used the consensus kiln count. We removed log night lights, because kilns burn through the night, and separately added degree of urbanisation, built-up surface and travel time [24]. For blood outcomes, we reweighted responders by the inverse of their predicted probability of a valid result, starting from the survey’s blood weight [11]. Finally, we compared February to March with June, the one month clearly after firing. Soil lead used blood sample weights, because soil was sampled only in blood-module households.

Analyses used complete cases; covariates were missing for 7 children. We used Stata 17 with version 15 syntax for estimation, and R 4 with sf and ggplot2 for exposure linkage and figures.

### Ethics

The icddr,b Ethical Review Committee approved the MICS 2025 protocol, including blood testing (PR-24113) [11]. Respondents gave verbal consent for interviews and written consent for blood collection [11]. We used anonymised public data, so no further approval was needed.

## Results

The analysis included 23,357 children under 5 years in 3,145 clusters; the other four clusters contributed no child interview (S2 Table). Of 17,124 children eligible for blood collection, 10,634 had a valid laboratory result and 10,622 a valid blood lead result. After excluding children with missing covariates or outcome, models included 23,341 children for cough, 23,331 for ARI symptoms and 10,618 for blood lead. Weighted estimates matched the survey report: diarrhoea 7.1%, ARI symptoms 2.9%, and blood lead of 5 µg/dL or more 38.5% (report 38.4%). The small difference arises because the report keeps all 10,661 children who gave blood in its denominator, including 27 without a valid laboratory result. Cough was reported for 28.4% of children (95% CI 27.6 to 29.2).

Three in four children (75.4%, 95% CI 73.6 to 77.1) lived within 5 km of at least one kiln (figure 2). One in five (20.3%, 18.7 to 22.0) had ten or more kilns within 5 km. For 30.3% (28.4 to 32.4), the nearest kiln was less than 2 km from the cluster point. Children had a mean of 6.9 kilns (6.4 to 7.4) within 5 km. Dense exposure was most common in Dhaka division (figure 1), where 37.0% (33.1 to 41.0) of children had ten or more kilns within 5 km. It was least common in Sylhet (3.4%, 1.6 to 7.2). It was more common in urban than rural areas (29.9%, 26.5 to 33.5, *vs* 16.4%, 14.6 to 18.3). It rose with wealth, from 10.1% (8.5 to 12.1) in the poorest quintile to 32.5% (29.3 to 35.8) in the richest (S2 Table).

**Figure 2.**
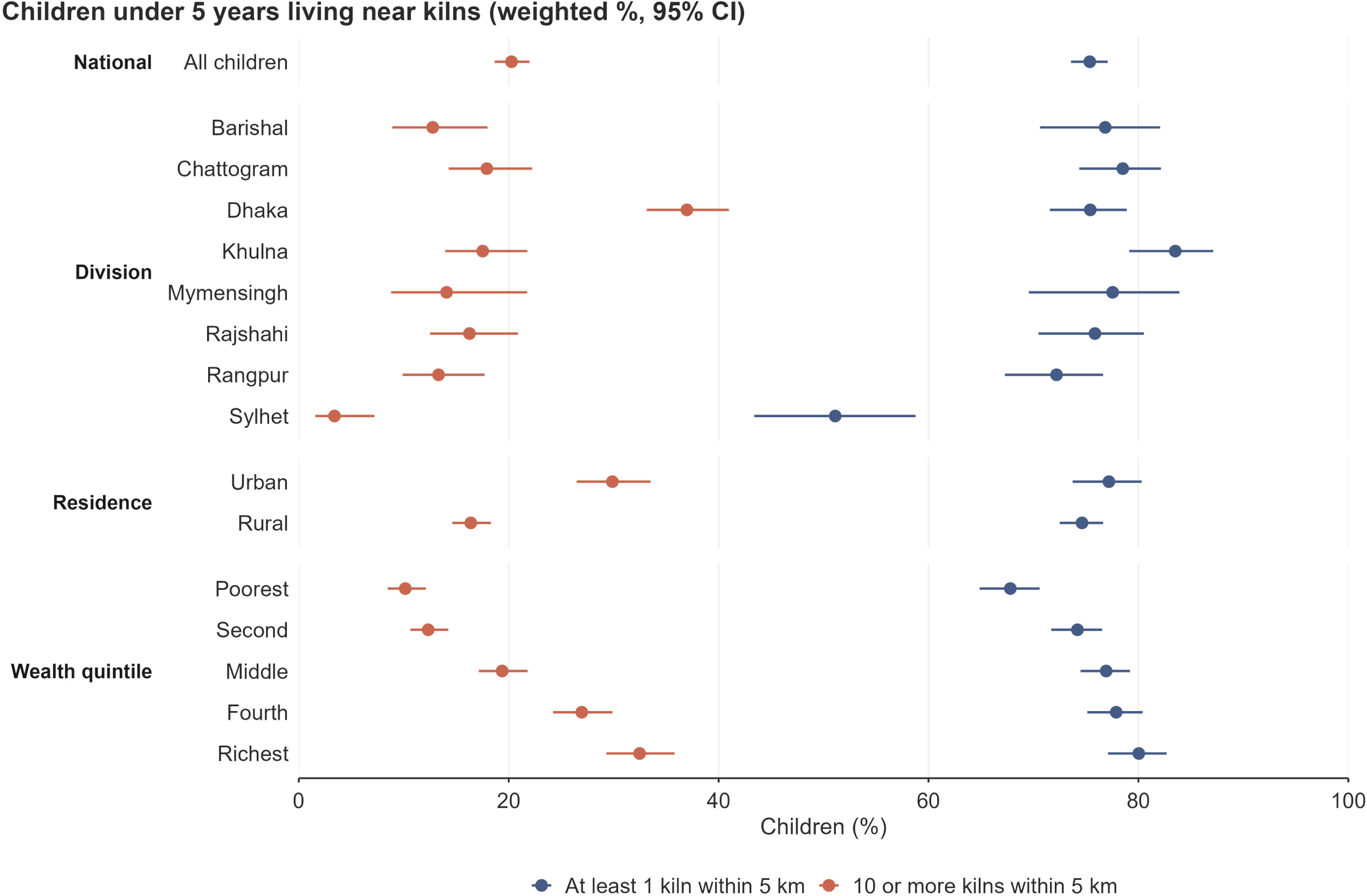
Children under 5 years living near brick kilns, by division, residence and wealth quintile. Weighted percentage of children with at least one kiln, and with ten or more kilns, within 5 km of the survey cluster, with 95% CIs. Exposure is measured at displaced cluster locations with the main kiln map.

Children in the most exposed group were more often urban, richer and in Dhaka division, and their households less often used solid fuel (table 1). Annual fine particulate matter in 2020 was higher in kiln-dense clusters (77.8 µg/m³ *vs* 71.4 µg/m³; table 1). District explained 26.7% of the variation in kiln counts between clusters (S2 Table), so most of the contrast lay within districts.

**Table 1.** Characteristics of children under 5 years by number of brick kilns within 5 km of the survey cluster, Bangladesh MICS 2025.

| Characteristic | None (n=6,338) | 1 to 4 (n=8,666) | 5 to 9 (n=4,395) | 10 or more (n=3,958) |
| --- | --- | --- | --- | --- |
| **Age (months)** |  |  |  |  |
| 0 to 11 | 19.4 | 19.8 | 18.1 | 19.6 |
| 12 to 23 | 19.6 | 20.7 | 21.4 | 22.3 |
| 24 to 35 | 21.1 | 21.1 | 21.3 | 20.4 |
| 36 to 47 | 21.3 | 19.3 | 18.9 | 19.1 |
| 48 to 59 | 18.5 | 19.1 | 20.3 | 18.6 |
| Sex: female | 48.7 | 48.3 | 47.7 | 47.4 |
| **Mother's education** |  |  |  |  |
| None or pre-primary | 7.8 | 5.7 | 5.3 | 4.8 |
| Primary | 21.8 | 17.4 | 16.7 | 15.3 |
| Secondary | 50.3 | 53.9 | 55.7 | 53.0 |
| Higher secondary or above | 20.2 | 22.9 | 22.3 | 26.9 |
| **Wealth quintile** |  |  |  |  |
| Poorest | 26.2 | 21.7 | 19.6 | 10.0 |
| Second | 20.7 | 21.6 | 23.2 | 12.0 |
| Middle | 18.7 | 20.2 | 22.2 | 19.1 |
| Fourth | 18.7 | 19.1 | 19.5 | 27.7 |
| Richest | 15.8 | 17.5 | 15.5 | 31.2 |
| Residence: urban | 26.6 | 25.4 | 23.4 | 42.4 |
| Cooking fuel: solid fuel | 77.2 | 79.9 | 82.3 | 55.6 |
| **Month of interview** |  |  |  |  |
| February to March (firing) | 30.5 | 37.3 | 36.8 | 34.9 |
| April (end of firing) | 32.6 | 25.7 | 25.0 | 23.3 |
| May to June (off season) | 37.0 | 37.0 | 38.3 | 41.8 |
| **Division** |  |  |  |  |
| Barishal | 5.0 | 5.8 | 6.8 | 3.3 |
| Chattogram | 20.6 | 26.3 | 25.5 | 20.9 |
| Dhaka | 24.1 | 18.0 | 14.7 | 44.0 |
| Khulna | 6.8 | 9.9 | 16.4 | 8.8 |
| Mymensingh | 7.5 | 9.5 | 9.4 | 5.7 |
| Rajshahi | 11.2 | 12.8 | 11.5 | 9.2 |
| Rangpur | 12.1 | 10.7 | 13.0 | 7.1 |
| Sylhet | 12.7 | 7.1 | 2.8 | 1.1 |
| **Place** |  |  |  |  |
| Distance to nearest kiln, km (mean) | 8.3 | 3.1 | 2.2 | 1.6 |
| Annual PM2.5 in 2020, µg/m<sup>3</sup> (mean) | 71.4 | 74.2 | 74.3 | 77.8 |
| **Outcomes** |  |  |  |  |
| Cough, last 2 weeks (%) | 26.7 | 28.8 | 29.4 | 28.8 |
| Symptoms of ARI, last 2 weeks (%) | 2.8 | 3.0 | 2.6 | 3.0 |
| Diarrhoea, last 2 weeks (%) | 6.8 | 6.9 | 8.1 | 6.6 |
| Blood lead 3.5 µg/dL or more (%) | 68.0 | 64.1 | 60.4 | 73.5 |
| Blood lead 5 µg/dL or more (%) | 40.0 | 33.7 | 33.1 | 50.8 |
| Blood lead, µg/dL (geometric mean) | 4.47 | 4.18 | 4.12 | 5.16 |

Weighted column percentages unless stated. n is the unweighted number of children with a completed under-five interview. Blood outcomes refer to children aged 12 to 59 months with a valid laboratory result and use blood sample weights. ARI=acute respiratory infection. PM2.5=fine particulate matter (MICS geospatial covariate).

Cough, ARI symptoms and diarrhoea differed little by kiln category (table 1). Compared with children with no kiln within 5 km, those with ten or more had an adjusted PR of 1.05 (0.96 to 1.16) for cough (figure 3). The PR for ARI symptoms was 1.21 (0.90 to 1.62). Neither outcome showed a trend across categories (P for trend 0.242 and 0.374). Diarrhoea, the negative control outcome, was also not associated (PR 0.97, 0.80 to 1.19). For ten or more kilns, the upper limit of the 95% CI rules out a cough excess of more than 16%. Per 10 additional kilns, the PR was 1.02 (0.99 to 1.05) for cough and 1.11 (1.01 to 1.21) for ARI symptoms. With the count top-coded at 55 kilns, the ARI estimate was 1.12 (1.00 to 1.25; S1 Table). A spline of the kiln count showed no association with cough (P=0.499) or ARI symptoms (P=0.153; S1 Fig). The ARI curve rose with kiln count, but its 95% CI excluded 1 only at 42 or more kilns, a level reached by 60 of 3,149 clusters.

**Figure 3.**
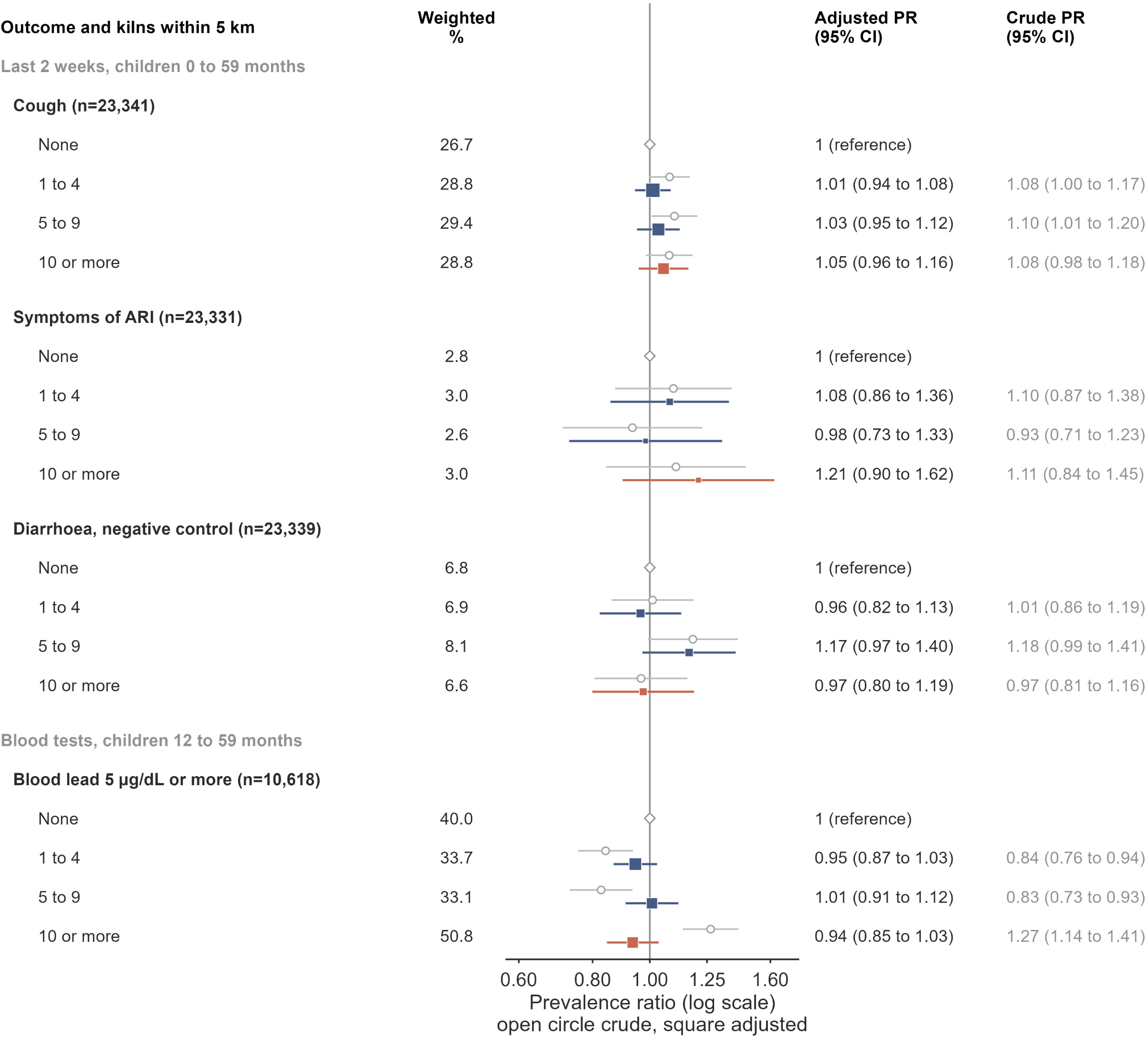
Crude and adjusted prevalence ratios for child outcomes by number of kilns within 5 km. Squares show adjusted prevalence ratios with 95% CIs, sized by precision; open circles show crude ratios. The reference is no kiln within 5 km. Adjusted models include child age, sex, mother’s education, wealth quintile, residence, cooking fuel, month of interview or blood draw, log population density, log night lights and district. Weighted % is the prevalence of each outcome in each kiln category. Diarrhoea is a negative control outcome. ARI=acute respiratory infection. PR=prevalence ratio.

Kiln density was not associated with cough at the peak of firing (figure 4). In February and March, the PR per 10 kilns was 1.00 (0.96 to 1.05), and in April 1.01 (0.96 to 1.07). In May and June, which span the end of firing and the month after it, the PR was 1.10 (1.04 to 1.17). The association differed across the three periods when kilns were counted per 10 (P for interaction 0.020) but not when grouped in categories (P=0.102). In June alone, the PR per 10 kilns was 1.14 (0.96 to 1.35), which did not differ clearly from February and March (P for interaction 0.425; S1 Table). For ten or more kilns, the cough PR was 0.95 (0.83 to 1.09) in February and March. It was 1.24 (1.04 to 1.49) in May and June, and 1.81 (1.28 to 2.56) in June alone (S1 and S3 Tables). ARI symptoms did not vary by period (P for interaction 0.534). Per 10 kilns, the PR was 1.10 (0.80 to 1.53) in February and March and 1.21 (1.06 to 1.39) in May and June.

**Figure 4.**
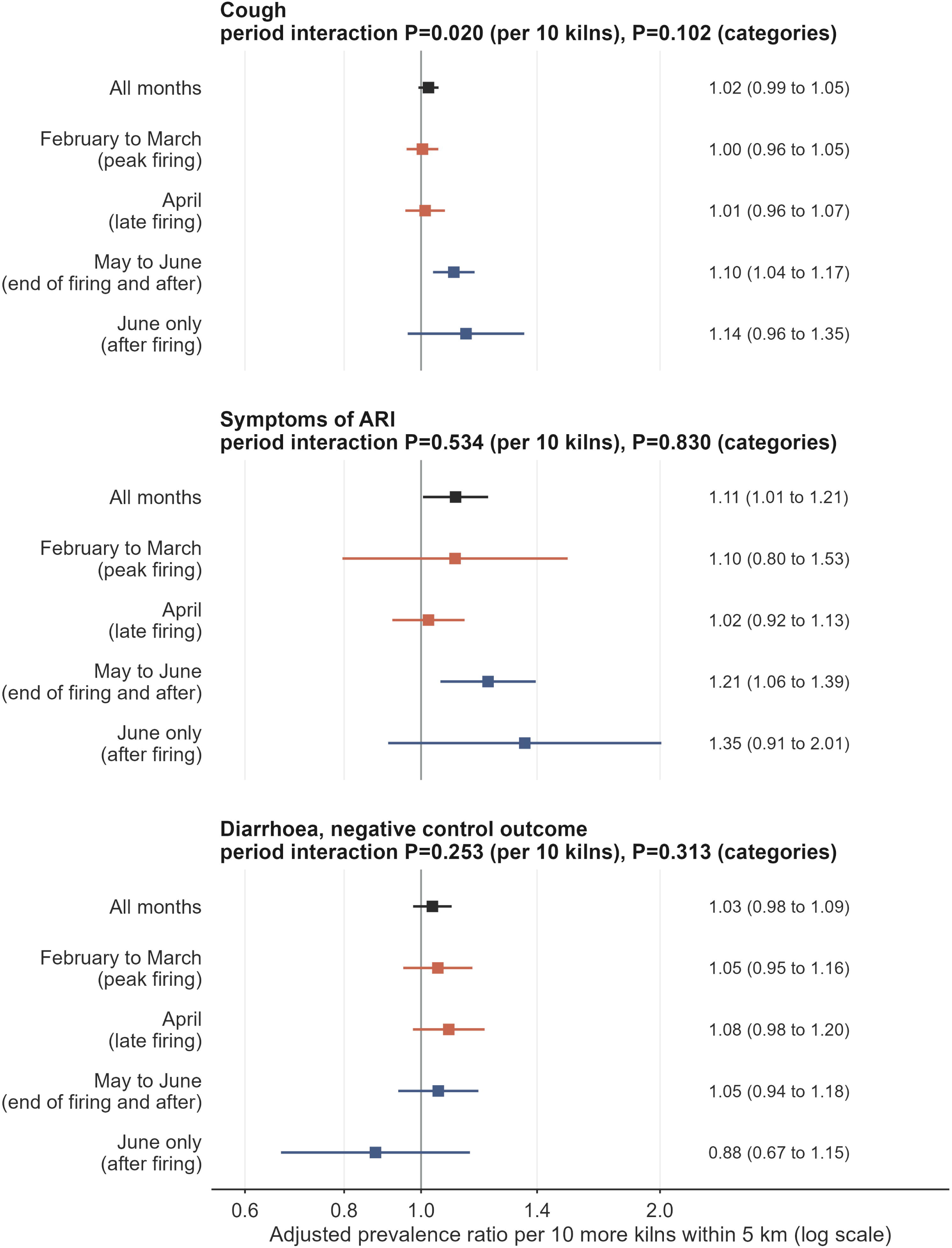
Kiln density and symptoms by month of interview. Adjusted prevalence ratios with 95% CIs per 10 additional kilns within 5 km, for all months and by period of interview. Kilns fire from about November to April or May, so February to March is the peak of firing, April is late in the season, May to June spans the end of firing and the month after it, and June alone is after firing. The interaction P values test whether the association differs across the three main periods, with kilns per 10 and as categories. The June-only row was added after review of a first draft. Models use the same adjustment as figure 3, including month of interview within periods.

Blood lead of 5 µg/dL or more was most common in the highest kiln category (50.8%), but lowest in the middle categories (33.7% and 33.1%; table 1). The crude PR for ten or more kilns was 1.27 (1.14 to 1.41). It fell to 1.10 (1.00 to 1.21) with child and household factors (S2 Fig and S4 Table). Adding urban setting, population density and night lights brought it to 1.04 (0.95 to 1.14). With district fixed effects the PR was 0.94 (0.85 to 1.03). Adding reported lead sources left it at 0.93 (0.84 to 1.03). The geometric mean ratio of blood lead was 0.98 (0.93 to 1.03; S5 Table), and a spline showed no association with blood lead (P=0.655; S1 Fig). The kiln association with blood lead varied by month of the blood draw (P for interaction 0.036; S3 Table), but estimates for ten or more kilns were below 1 in every period. Lead has a mean life of about 35 days in blood [27], so this contrast says little about firing.

Blood response was lower among eligible children with ten or more kilns within 5 km (55.2%, 53.0 to 57.5) than among those with none (62.6%, 60.7 to 64.6; S1 Table). Reweighting for non-response left the estimates for ten or more kilns unchanged (table 2). The PR for blood lead of 5 µg/dL or more was 0.95 (0.86 to 1.05).

**Table 2.** Adjusted prevalence ratios for the highest versus the lowest kiln exposure, main model and sensitivity analyses.

| Analysis | Cough | Symptoms of ARI | Blood lead 5 µg/dL or more |
| --- | --- | --- | --- |
| Main model: 10 or more kilns within 5 km vs none | 1.05 (0.96 to 1.16) | 1.21 (0.90 to 1.62) | 0.94 (0.85 to 1.03) |
| Nearest kiln under 2 km vs 10 km or more | 1.28 (1.11 to 1.48) | 1.22 (0.77 to 1.93) | 1.00 (0.85 to 1.17) |
| 30 or more kilns within 10 km vs none | 1.35 (1.15 to 1.57) | 1.27 (0.76 to 2.11) | 1.02 (0.85 to 1.23) |
| Six divisions mapped by both sources: main kiln map | 1.01 (0.90 to 1.14) | 0.96 (0.66 to 1.40) | 1.01 (0.90 to 1.13) |
| Six divisions mapped by both sources: second kiln map (APAD) | 0.98 (0.86 to 1.12) | 0.97 (0.61 to 1.55) | 0.91 (0.78 to 1.05) |
| Urban clusters only (displacement up to 2 km) | 1.07 (0.90 to 1.27) | 2.37 (1.33 to 4.21) | 0.98 (0.87 to 1.11) |
| Excluding clusters within 5 km of the land border | 1.06 (0.96 to 1.17) | 1.25 (0.93 to 1.68) | 0.95 (0.86 to 1.05) |
| Kilns found by both maps, six divisions | 1.06 (0.93 to 1.20) | 1.11 (0.75 to 1.65) | 1.06 (0.95 to 1.19) |
| Without log night lights | 1.04 (0.95 to 1.15) | 1.22 (0.91 to 1.64) | 0.98 (0.88 to 1.08) |
| Adding urbanisation class, built-up surface and travel time | 1.05 (0.95 to 1.16) | 1.25 (0.92 to 1.68) | 0.93 (0.84 to 1.03) |
| Weights for blood non-response (blood outcomes only) | Not applicable | Not applicable | 0.95 (0.86 to 1.05) |

Prevalence ratios (95% CI) from survey-weighted modified Poisson models adjusted for child age, sex, mother's education, wealth quintile, residence, cooking fuel, month of interview or blood draw, log population density, log night lights and district. The second kiln map (APAD) covers Barishal, Dhaka, Khulna, Mymensingh, Rajshahi and Rangpur divisions only, so both maps are compared within these six divisions. The urban model omits the residence term. Rows below the urban row were added after review of the first draft and are post hoc. Kilns found by both maps: main-map kilns with a second-map kiln within 500 m. Weights for blood non-response: child weight times the number of children aged 12 to 59 months in the household, divided by the predicted probability of a valid blood result among eligible children, truncated at the 99th centile.

Household soil lead had a geometric mean of 21.5 mg/kg in 10,550 households (S6 Table). Soil lead was 11% higher in the highest kiln category before adjustment (ratio 1.11, 1.06 to 1.17) but not after it (0.99, 0.95 to 1.04; S6 Table). Among exploratory metals, arsenic and mercury showed no association. Blood cadmium was lower with more kilns (geometric mean ratio for ten or more kilns 0.84, 0.77 to 0.91; P for trend <0.001). This persisted without night lights (0.85, 0.79 to 0.92) and with added place covariates (0.84, 0.77 to 0.91; S1 Table).

The second kiln map gave similar results within the six divisions it covers, as did the count of kilns found by both maps (table 2). Three further checks changed no estimate for ten or more kilns by more than 0.05: excluding clusters within 5 km of the land border, removing night lights and adding place covariates (table 2 and S1 Table). Blood lead remained unrelated to kilns with every exposure measure (S7 Table).

Cough was more common in all groups with a kiln within 10 km than in the 5.2% of children with none (table 2 and S7 Table). The PR was 1.26 (1.10 to 1.44) for 1 to 9 kilns, 1.33 (1.15 to 1.53) for 10 to 29 and 1.35 (1.15 to 1.57) for 30 or more (P for trend 0.004). Most of this gradient was the step from none to any kiln, and nearest-kiln distance gave similar PRs of about 1.3 in every band under 10 km (S7 Table). The reference clusters lay mainly in Chattogram, Sylhet and Rangpur (S7 Table). The excess remained when clusters within 10 km of the land border were excluded (PR 1.43, 1.16 to 1.77, for 30 or more kilns; S1 Table), so missing kilns across the border do not explain it.

In urban clusters, ten or more kilns were associated with ARI symptoms (PR 2.37, 1.33 to 4.21; table 2), based on 49 and 34 cases in the highest and reference groups (S7 Table). This was one of many subgroup estimates and was not planned as a primary analysis.

A second random displacement changed exposure only moderately. The 5 km kiln count at the re-displaced point correlated with the published count (Spearman 0.86; 2.5th to 97.5th percentile of 100 runs 0.85 to 0.87; S8 Table). The Spearman correlation was higher in urban clusters (0.94) than in rural ones (0.82). Three in four clusters stayed in the same category (85% urban, 71% rural). Within districts, the correlation was 0.85 (0.80 to 0.89) and the slope of the re-displaced on the published count was 0.86 (0.78 to 0.92). Displacement would therefore shrink a log-linear per-kiln association by about 14%. The two kiln maps agreed less well than the displaced points. They shared 47% of the main map’s kilns within 500 m. In the six divisions both maps cover, their 5 km counts had a Spearman correlation of 0.56 (S8 Table).

## Discussion

Three in four young children in Bangladesh live within 5 km of a brick kiln, and one in five lives among ten or more. Within the resolution of displaced survey coordinates, kiln density was not associated with blood lead or household soil lead once place was held fixed. These results held with a spline of kiln count, a second kiln map, kilns confirmed by both maps, exclusion of border clusters and reweighting for blood non-response. Associations with respiratory symptoms were weak and inconsistent. ARI symptoms rose slightly per 10 kilns but not across categories or in a spline. Cough was more common with any kiln within 10 km but was not associated with kiln density at the peak of firing. Together these patterns are more consistent with confounding by place than with local harm from kiln smoke, although they cannot exclude a small effect.

Our exposure estimates agree with the satellite study that found at least 12% of people within 1 km of a kiln and 95% within 10 km [1]. We add that exposure is greatest for urban and richer children and in Dhaka division. This runs against the usual pattern of pollution falling hardest on the poor [7]. In Mexico, by contrast, artisanal kilns sit within marginalised communities [28].

Kilns ring growing cities, where demand for bricks is highest [1,3]. The same pattern explains why kiln density first appeared to track blood lead. Blood lead is higher in urban and wealthier households and in Dhaka division [11], and kilns concentrate there as well. Once district and urban setting were held fixed, the lead excess disappeared.

The null result for lead fits what is known about lead sources. Informal battery recycling, electronic waste and lead-adulterated spices are major sources in low-income and middle-income countries [29], and lead chromate in turmeric has been a primary source in Bangladesh [30]. The survey report itself found far higher blood lead in children living near industrial areas or lead-related activity than in other children [11]. Soil within 200 m of kilns in Natore had lead above regional reference values, but not in relation to distance from the chimney [10]. Children living at kiln sites in Pakistan had nickel, cadmium and chromium in their blood [12]. Those studies sampled within a few hundred metres of kilns or among families living on kiln premises. Our design compared neighbourhoods several kilometres apart. Household soil lead did not rise with kiln density after adjustment, which suggests little kiln lead deposition at this scale. We cannot exclude effects for children who live or work on kiln premises.

For respiratory symptoms, the field study by Brooks and colleagues found raised odds of symptoms in adults within 2 km downwind during the production season [2]. Its estimate for children under 5 years was imprecise, with odds ratio 2.1 (0.7 to 6.0) [2]. We found no association with cough at the peak of firing. Brooks and colleagues used exact homes, wind direction and a 2 km downwind zone, while our coordinates blur location by up to 5 km. In Dhaka, the link between daily fine particles and childhood pneumonia was stronger on days with a larger kiln share [9]. Such day-to-day regional exposure is shared by children across a district, so our within-district contrast cannot capture it. In Nepal, workers and family members living at kilns had high personal exposures and signs of small-airway disease, although few reported respiratory symptoms [31]. Such on-site exposure is far closer than our neighbourhood contrast can resolve.

The season contrast is informative but not decisive. If kiln smoke raised cough, the association should be present while kilns burn. It was absent in February, March and April and appeared in May and June, as firing ended. Two points limit this test. Kilns fire from about November to April or May [1,26], so only June is clearly after firing, and the June estimate was imprecise and did not differ clearly from the firing months. In addition, the survey report cautions that two-week symptom prevalence varies with season, local outbreaks and the timing of team visits [11], and these may differ between crowded peri-urban areas and rural ones. Confounding that changes with season, or a lagged effect of months of exposure, could produce the same pattern. The absence of any association at the peak of firing is the more robust finding. ARI symptoms did not vary by period and give no support either way.

The cough excess near any kiln within 10 km also points to place. Children with any kiln within 10 km had more cough than the few with none, with little further rise across distance bands or kiln counts. The comparison rests on few remote clusters, mostly in Sylhet, Chattogram and Rangpur divisions, which likely differ in ways we could not measure. The excess was not explained by kilns missing across the border.

The inverse association of kilns with blood cadmium is a warning about residual confounding. Kilns have no known way to lower cadmium, yet the association survived district fixed effects and finer place covariates. Rice is a major source of dietary cadmium in Bangladesh [32,33], and diets may differ between urban and rural areas, and kiln-dense areas are more urban. Residual confounding of this kind could bias the other estimates in either direction, including toward the null for lead.

The urban result for ARI symptoms needs caution. It rests on few cases and was one of many comparisons. It does fit the idea that better location precision reveals effects that displacement hides [20]. Urban clusters are displaced by at most 2 km, and their exposure was the most stable in our simulation. This finding should be tested in studies with exact addresses.

Our results do not show that kilns are harmless. Kiln emissions spread far and form secondary particles, so much of their burden falls on whole regions [6]. A within-district contrast removes that shared regional exposure by design. Our results speak to the extra risk of living closer to more kilns, not to the total burden of the brick industry. Better operating practices in zigzag kilns cut fuel use and emissions without new capital investment in a recent trial in Bangladesh [26]. Such measures would reduce regional pollution whatever the local gradient.

This study has several strengths. It used a national probability sample with venous blood measures. We reproduced the survey report, used two independent kiln maps and checked confounding with a negative control outcome, a negative control period and an unplanned bias marker in cadmium. We quantified the stability of exposure under displacement, within districts as well as overall, and tested the main results against border truncation, map error and blood non-response.

Several limitations apply. First, displaced coordinates blur location. Within districts, a second displacement would shrink a per-kiln association only modestly, so displacement alone is unlikely to hide a large effect at the 5 km scale. It does hide effects within 1 to 2 km. Non-differential error of this kind usually biases estimates toward the null, although this is not guaranteed for the middle categories of a grouped exposure. Second, the main kiln map is from 2018 to 2019, six years before the survey. Kilns open, close and convert, and the two maps agreed only moderately; the second map’s Bangladesh layer also retains false positives [17].

Agreement of two error-prone maps on a null result does not rule out attenuation, although the consensus count, which keeps kilns found by both maps, gave the same result. Third, the main map stops at the border; excluding border clusters did not change the results. Fourth, we lacked wind direction, kiln operating status and kiln size, so our count is a crude proxy for exposure.

Other limitations concern the outcomes and the analysis. Fifth, symptoms were reported by mothers over 2 weeks and may be misclassified. The survey report advises that these data describe period prevalence rather than the epidemiology of the illnesses [11]. Sixth, a third of eligible children gave no blood, and response was lower in kiln-dense areas. Reweighting by measured factors did not change the results, but bias remains possible if refusal depended on blood lead itself. Seventh, we limited blood outcomes to lead and did not analyse haemoglobin, which the survey also measured. Eighth, we produced many estimates, and some analyses were added after review of a first draft; isolated results near P=0.05 should be read as hypothesis-generating. Finally, the cross-sectional design cannot establish timing, and the cadmium result shows that residual confounding by place remains.

## Conclusions

Most young children in Bangladesh live near brick kilns, and the richest and most urban children live among the most. At the resolution of national survey data, children in kiln-dense areas did not have more blood lead or soil lead. Respiratory associations were weak and did not appear at the peak of firing, which fits confounding by place better than an acute effect of kiln smoke. Lead control should keep its focus on proven sources [29,30], while studies with exact locations should measure the near-field and regional burden of kiln smoke.

## Data Availability

MICS 2025 data and cluster coordinates are available from UNICEF (https://mics.unicef.org) on registration. The kiln coordinates are publicly available from the Harvard Dataverse (doi:10.7910/DVN/HVGW8L) and the APAD asset data repository.

## Declarations

### Contributors

SMAUH conceived the study and designed the methodology. He curated the data, performed the formal analysis, and created the visualizations. He also wrote the original draft, revised the manuscript, and managed the project. MKH contributed to data curation, wrote part of the original draft, and reviewed and edited the manuscript. MKI, MMS, and SMRH contributed to interpretation of the results and reviewed the manuscript. SAJ contributed to interpretation of the results and reviewed and edited the manuscript. KUB supervised the study, validated the results, and reviewed and edited the manuscript. All authors read and approved the final manuscript.

## Data sharing

Aggregate results are in the tables and supplementary material.

## Acknowledgments

We thank the Bangladesh Bureau of Statistics, UNICEF, icddr,b and the survey participants.

---

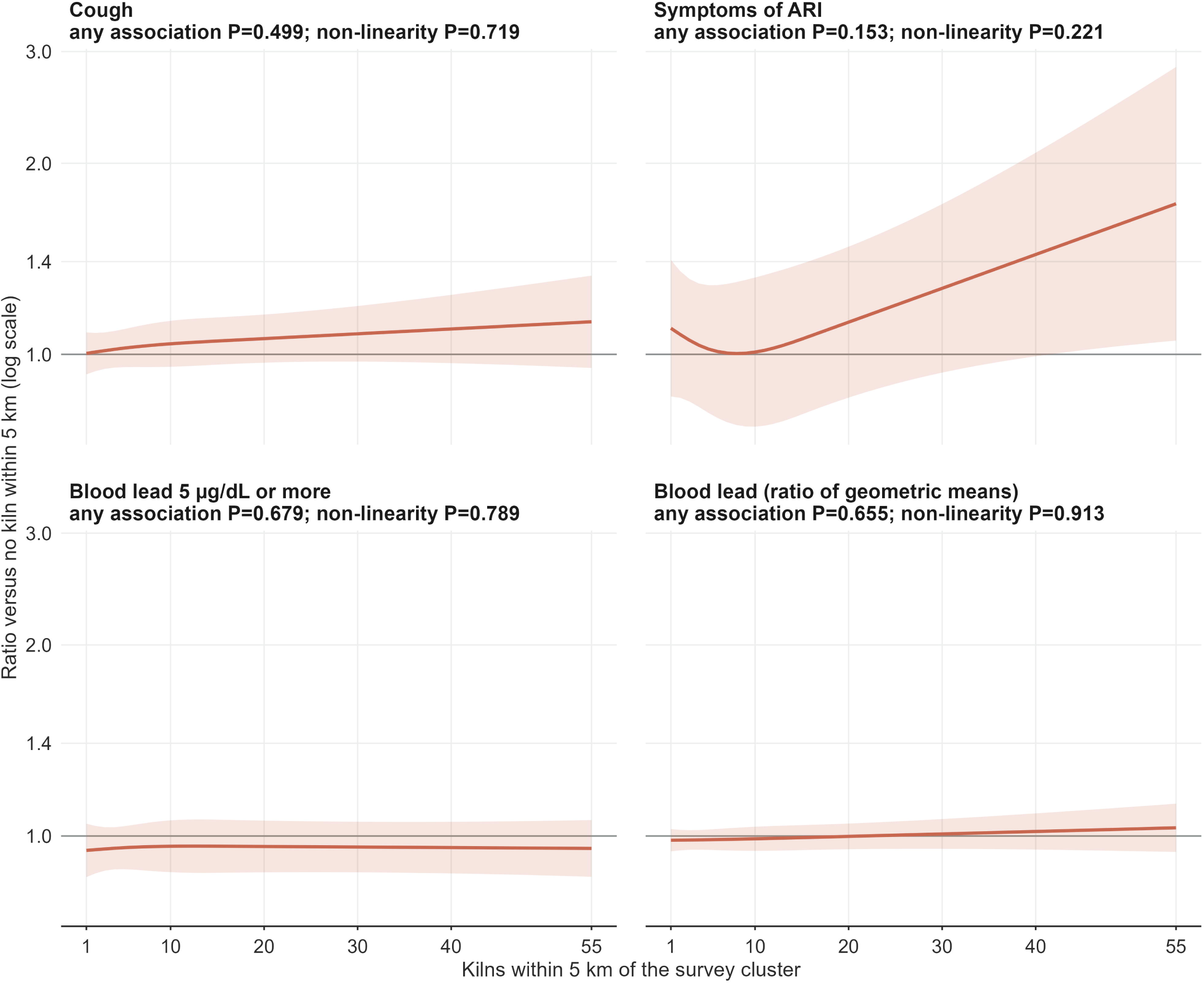

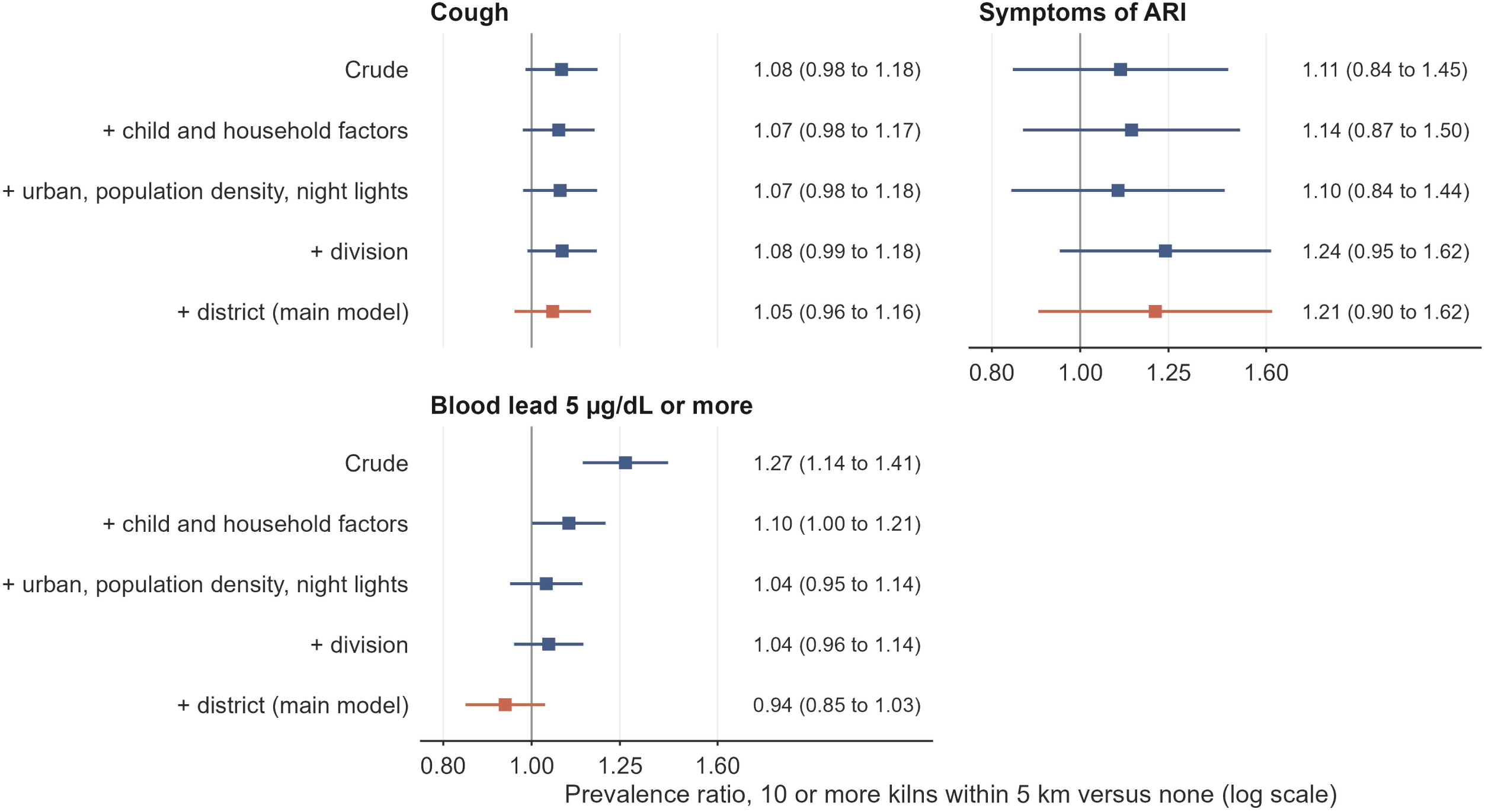

