## Supplementary material for "Brick kilns, child respiratory symptoms and blood lead in Bangladesh: a national geospatial analysis of the 2025 Multiple Indicator Cluster Survey": Supplement_Draft_v7.docx

### S1 Table. Analyses added after review of the first draft (post hoc)

| Analysis | Outcome | Level | Estimate | P for trend | n |
| --- | --- | --- | --- | --- | --- |
| Excluding clusters within 5 km of the land border | Cough | 1 to 4 | 1.01 (0.94 to 1.08) | 0.187 | 21,609 |
| Excluding clusters within 5 km of the land border | Cough | 5 to 9 | 1.04 (0.96 to 1.13) | 0.187 | 21,609 |
| Excluding clusters within 5 km of the land border | Cough | 10 or more | 1.06 (0.96 to 1.17) | 0.187 | 21,609 |
| Kilns within 10 km, excluding clusters within 10 km of the land border | Cough | 1 to 9 | 1.36 (1.12 to 1.66) | 0.018 | 19,972 |
| Kilns within 10 km, excluding clusters within 10 km of the land border | Cough | 10 to 29 | 1.43 (1.17 to 1.75) | 0.018 | 19,972 |
| Kilns within 10 km, excluding clusters within 10 km of the land border | Cough | 30 or more | 1.43 (1.16 to 1.77) | 0.018 | 19,972 |
| Excluding clusters within 5 km of the land border | Symptoms of ARI | 1 to 4 | 1.08 (0.85 to 1.37) | 0.266 | 21,599 |
| Excluding clusters within 5 km of the land border | Symptoms of ARI | 5 to 9 | 0.99 (0.73 to 1.34) | 0.266 | 21,599 |
| Excluding clusters within 5 km of the land border | Symptoms of ARI | 10 or more | 1.25 (0.93 to 1.68) | 0.266 | 21,599 |
| Kilns within 10 km, excluding clusters within 10 km of the land border | Symptoms of ARI | 1 to 9 | 0.89 (0.52 to 1.51) | 0.716 | 19,962 |
| Kilns within 10 km, excluding clusters within 10 km of the land border | Symptoms of ARI | 10 to 29 | 1.03 (0.58 to 1.83) | 0.716 | 19,962 |
| Kilns within 10 km, excluding clusters within 10 km of the land border | Symptoms of ARI | 30 or more | 0.95 (0.52 to 1.74) | 0.716 | 19,962 |
| Excluding clusters within 5 km of the land border | Blood lead 5 µg/dL or more | 1 to 4 | 0.92 (0.85 to 1.01) | 0.686 | 9,755 |
| Excluding clusters within 5 km of the land border | Blood lead 5 µg/dL or more | 5 to 9 | 1.02 (0.92 to 1.13) | 0.686 | 9,755 |
| Excluding clusters within 5 km of the land border | Blood lead 5 µg/dL or more | 10 or more | 0.95 (0.86 to 1.05) | 0.686 | 9,755 |
| Kilns within 10 km, excluding clusters within 10 km of the land border | Blood lead 5 µg/dL or more | 1 to 9 | 0.94 (0.78 to 1.15) | 0.574 | 8,968 |
| Kilns within 10 km, excluding clusters within 10 km of the land border | Blood lead 5 µg/dL or more | 10 to 29 | 0.96 (0.79 to 1.18) | 0.574 | 8,968 |
| Kilns within 10 km, excluding clusters within 10 km of the land border | Blood lead 5 µg/dL or more | 30 or more | 0.99 (0.80 to 1.23) | 0.574 | 8,968 |
| Kilns found by both maps, six divisions | Cough | 1 to 4 | 1.00 (0.92 to 1.09) | 0.438 | 16,545 |
| Kilns found by both maps, six divisions | Cough | 5 to 9 | 1.01 (0.91 to 1.13) | 0.438 | 16,545 |
| Kilns found by both maps, six divisions | Cough | 10 or more | 1.06 (0.93 to 1.20) | 0.438 | 16,545 |
| Kilns found by both maps, six divisions | Symptoms of ARI | 1 to 4 | 0.96 (0.71 to 1.30) | 0.986 | 16,546 |
| Kilns found by both maps, six divisions | Symptoms of ARI | 5 to 9 | 0.81 (0.56 to 1.17) | 0.986 | 16,546 |
| Kilns found by both maps, six divisions | Symptoms of ARI | 10 or more | 1.11 (0.75 to 1.65) | 0.986 | 16,546 |
| Kilns found by both maps, six divisions | Blood lead 5 µg/dL or more | 1 to 4 | 1.11 (1.01 to 1.21) | 0.328 | 7,820 |
| Kilns found by both maps, six divisions | Blood lead 5 µg/dL or more | 5 to 9 | 1.03 (0.89 to 1.18) | 0.328 | 7,820 |
| Kilns found by both maps, six divisions | Blood lead 5 µg/dL or more | 10 or more | 1.06 (0.95 to 1.19) | 0.328 | 7,820 |
| Without log night lights | Cough | 1 to 4 | 1.01 (0.94 to 1.09) | 0.324 | 23,341 |
| Without log night lights | Cough | 5 to 9 | 1.03 (0.95 to 1.12) | 0.324 | 23,341 |
| Without log night lights | Cough | 10 or more | 1.04 (0.95 to 1.15) | 0.324 | 23,341 |
| Adding urbanisation class, built-up surface and travel time | Cough | 1 to 4 | 1.01 (0.94 to 1.08) | 0.257 | 23,341 |
| Adding urbanisation class, built-up surface and travel time | Cough | 5 to 9 | 1.03 (0.95 to 1.12) | 0.257 | 23,341 |
| Adding urbanisation class, built-up surface and travel time | Cough | 10 or more | 1.05 (0.95 to 1.16) | 0.257 | 23,341 |
| Without log night lights | Symptoms of ARI | 1 to 4 | 1.08 (0.86 to 1.36) | 0.326 | 23,331 |
| Without log night lights | Symptoms of ARI | 5 to 9 | 0.99 (0.73 to 1.33) | 0.326 | 23,331 |
| Without log night lights | Symptoms of ARI | 10 or more | 1.22 (0.91 to 1.64) | 0.326 | 23,331 |
| Adding urbanisation class, built-up surface and travel time | Symptoms of ARI | 1 to 4 | 1.13 (0.89 to 1.43) | 0.282 | 23,331 |
| Adding urbanisation class, built-up surface and travel time | Symptoms of ARI | 5 to 9 | 1.03 (0.76 to 1.40) | 0.282 | 23,331 |
| Adding urbanisation class, built-up surface and travel time | Symptoms of ARI | 10 or more | 1.25 (0.92 to 1.68) | 0.282 | 23,331 |
| Without log night lights | Blood lead 5 µg/dL or more | 1 to 4 | 0.94 (0.86 to 1.03) | 0.998 | 10,618 |
| Without log night lights | Blood lead 5 µg/dL or more | 5 to 9 | 1.02 (0.92 to 1.13) | 0.998 | 10,618 |
| Without log night lights | Blood lead 5 µg/dL or more | 10 or more | 0.98 (0.88 to 1.08) | 0.998 | 10,618 |
| Adding urbanisation class, built-up surface and travel time | Blood lead 5 µg/dL or more | 1 to 4 | 0.94 (0.86 to 1.02) | 0.379 | 10,618 |
| Adding urbanisation class, built-up surface and travel time | Blood lead 5 µg/dL or more | 5 to 9 | 1.00 (0.91 to 1.12) | 0.379 | 10,618 |
| Adding urbanisation class, built-up surface and travel time | Blood lead 5 µg/dL or more | 10 or more | 0.93 (0.84 to 1.03) | 0.379 | 10,618 |
| Without log night lights | Log blood cadmium (difference) | 1 to 4 | -0.08 (-0.13 to -0.03) | <0.001 | 10,619 |
| Without log night lights | Log blood cadmium (difference) | 5 to 9 | -0.08 (-0.14 to -0.01) | <0.001 | 10,619 |
| Without log night lights | Log blood cadmium (difference) | 10 or more | -0.16 (-0.24 to -0.08) | <0.001 | 10,619 |
| Adding urbanisation class, built-up surface and travel time | Log blood cadmium (difference) | 1 to 4 | -0.08 (-0.13 to -0.03) | <0.001 | 10,619 |
| Adding urbanisation class, built-up surface and travel time | Log blood cadmium (difference) | 5 to 9 | -0.09 (-0.16 to -0.02) | <0.001 | 10,619 |
| Adding urbanisation class, built-up surface and travel time | Log blood cadmium (difference) | 10 or more | -0.18 (-0.26 to -0.10) | <0.001 | 10,619 |
| Weights for blood non-response | Blood lead 5 µg/dL or more | 1 to 4 | 0.95 (0.87 to 1.03) | 0.577 | 10,618 |
| Weights for blood non-response | Blood lead 5 µg/dL or more | 5 to 9 | 1.01 (0.91 to 1.12) | 0.577 | 10,618 |
| Weights for blood non-response | Blood lead 5 µg/dL or more | 10 or more | 0.95 (0.86 to 1.05) | 0.577 | 10,618 |
| Weights for blood non-response | Log blood lead (difference) | 1 to 4 | -0.02 (-0.05 to 0.02) | 0.815 | 10,618 |
| Weights for blood non-response | Log blood lead (difference) | 5 to 9 | 0.01 (-0.04 to 0.05) | 0.815 | 10,618 |
| Weights for blood non-response | Log blood lead (difference) | 10 or more | -0.02 (-0.07 to 0.04) | 0.815 | 10,618 |
| February to March interviews only | Cough | 1 to 4 | 0.95 (0.85 to 1.05) | 0.627 | 8,075 |
| February to March interviews only | Cough | 5 to 9 | 0.98 (0.86 to 1.11) | 0.627 | 8,075 |
| February to March interviews only | Cough | 10 or more | 0.95 (0.83 to 1.09) | 0.627 | 8,075 |
| June interviews only | Cough | 1 to 4 | 1.07 (0.84 to 1.37) | <0.001 | 2,352 |
| June interviews only | Cough | 5 to 9 | 1.19 (0.88 to 1.60) | <0.001 | 2,352 |
| June interviews only | Cough | 10 or more | 1.81 (1.28 to 2.56) | <0.001 | 2,352 |
| February to March interviews only | Symptoms of ARI | 1 to 4 | 1.09 (0.77 to 1.55) | 0.592 | 8,073 |
| February to March interviews only | Symptoms of ARI | 5 to 9 | 0.95 (0.62 to 1.44) | 0.592 | 8,073 |
| February to March interviews only | Symptoms of ARI | 10 or more | 1.26 (0.79 to 2.02) | 0.592 | 8,073 |
| June interviews only | Symptoms of ARI | 1 to 4 | 1.26 (0.54 to 2.96) | 0.444 | 2,352 |
| June interviews only | Symptoms of ARI | 5 to 9 | 0.85 (0.21 to 3.42) | 0.444 | 2,352 |
| June interviews only | Symptoms of ARI | 10 or more | 1.69 (0.45 to 6.37) | 0.444 | 2,352 |
| February to March interviews only | Diarrhoea (negative control outcome) | 1 to 4 | 1.04 (0.80 to 1.37) | 0.080 | 8,076 |
| February to March interviews only | Diarrhoea (negative control outcome) | 5 to 9 | 1.39 (1.05 to 1.85) | 0.080 | 8,076 |
| February to March interviews only | Diarrhoea (negative control outcome) | 10 or more | 1.23 (0.86 to 1.74) | 0.080 | 8,076 |
| June interviews only | Diarrhoea (negative control outcome) | 1 to 4 | 0.54 (0.26 to 1.10) | 0.198 | 2,352 |
| June interviews only | Diarrhoea (negative control outcome) | 5 to 9 | 0.82 (0.38 to 1.78) | 0.198 | 2,352 |
| June interviews only | Diarrhoea (negative control outcome) | 10 or more | 0.45 (0.20 to 1.01) | 0.198 | 2,352 |

| Analysis | Outcome | Period | Estimate | P | n |
| --- | --- | --- | --- | --- | --- |
| Per 10 kilns, count top-coded at 55 | Cough | All months | 1.03 (1.00 to 1.07) | 0.064 | 23,341 |
| Per 10 kilns, count top-coded at 55 | Symptoms of ARI | All months | 1.12 (1.00 to 1.25) | 0.059 | 23,331 |
| Per 10 kilns, count top-coded at 55 | Blood lead 5 µg/dL or more | All months | 0.99 (0.97 to 1.02) | 0.496 | 10,618 |
| Per 10 kilns, count top-coded at 55 | Log blood lead (ratio of geometric means) | All months | 1.01 (0.99 to 1.03) | 0.333 | 10,618 |
| Per 10 kilns, by period | Cough | February to March | 1.00 (0.96 to 1.05) | 0.867 | 8,075 |
| Per 10 kilns, by period | Cough | June | 1.14 (0.96 to 1.35) | 0.131 | 2,352 |
| Per 10 kilns, by period | Symptoms of ARI | February to March | 1.10 (0.80 to 1.53) | 0.554 | 8,073 |
| Per 10 kilns, by period | Symptoms of ARI | June | 1.35 (0.91 to 2.01) | 0.137 | 2,352 |
| Per 10 kilns, by period | Diarrhoea (negative control outcome) | February to March | 1.05 (0.95 to 1.16) | 0.341 | 8,076 |
| Per 10 kilns, by period | Diarrhoea (negative control outcome) | June | 0.88 (0.67 to 1.15) | 0.344 | 2,352 |

| Outcome | Test | F | P |
| --- | --- | --- | --- |
| Cough | Spline: non-linearity among exposed clusters | 0.13 | 0.719 |
| Cough | Spline: any association | 0.79 | 0.499 |
| Symptoms of ARI | Spline: non-linearity among exposed clusters | 1.50 | 0.221 |
| Symptoms of ARI | Spline: any association | 1.76 | 0.153 |
| Blood lead 5 µg/dL or more | Spline: non-linearity among exposed clusters | 0.07 | 0.789 |
| Blood lead 5 µg/dL or more | Spline: any association | 0.50 | 0.679 |
| Log blood lead | Spline: non-linearity among exposed clusters | 0.01 | 0.913 |
| Log blood lead | Spline: any association | 0.54 | 0.655 |
| Cough | Per 10 kilns by period (February to March vs June) | 0.64 | 0.425 |
| Symptoms of ARI | Per 10 kilns by period (February to March vs June) | 0.68 | 0.410 |
| Diarrhoea (negative control outcome) | Per 10 kilns by period (February to March vs June) | 0.54 | 0.463 |

| Kilns within 5 km | Eligible children | Valid blood result | Weighted response, % (95% CI) |
| --- | --- | --- | --- |
| None | 4,622 | 2,944 | 62.6 (60.7 to 64.6) |
| 1 to 4 | 6,268 | 3,948 | 61.8 (60.1 to 63.6) |
| 5 to 9 | 3,286 | 2,084 | 61.9 (59.5 to 64.3) |
| 10 or more | 2,948 | 1,658 | 55.2 (53.0 to 57.5) |

Prevalence ratios (95% CI) unless stated; differences for log cadmium and log blood lead. All models use the main adjustment set. Land border: Bangladesh outline within 3 km of India or Myanmar in a coarse world map. Spline: restricted cubic spline of the 5 km count among clusters with at least one kiln (knots at the 10th, 50th and 90th centiles of non-zero counts), with an indicator for any kiln; curves are in S1 Fig. Response is a valid laboratory result among children eligible for the blood module.

### S2 Table. Analytic sample, and children under 5 years living near brick kilns by division, residence and wealth quintile

| Step | n |
| --- | --- |
| Children under 5 years with a completed interview | 23,357 |
| Clusters with at least one interviewed child | 3,145 |
| Analysed for cough (complete covariates and outcome) | 23,341 |
| Analysed for symptoms of ARI | 23,331 |
| Eligible for blood collection (one child aged 12 to 59 months per household) | 17,124 |
| Valid laboratory result | 10,634 |
| Valid blood lead result | 10,622 |
| Analysed for blood lead 5 µg/dL or more | 10,618 |
| Household soil lead result | 10,550 |
| Analysed for soil lead | 10,546 |

Unweighted counts. The survey report lists 24,680 children under 5 years and 17,132 children eligible for blood collection; the eligible count here is restricted to children with a completed under-five interview.

| Group | Level | Unweighted n | At least 1 kiln within 5 km, % | 10 or more kilns within 5 km, % | Nearest kiln under 2 km, % | Mean kilns within 5 km |
| --- | --- | --- | --- | --- | --- | --- |
| National | All | 23,357 | 75.4 (73.6 to 77.1) | 20.3 (18.7 to 22.0) | 30.3 (28.4 to 32.4) | 6.9 (6.4 to 7.4) |
| Division | Barishal | 1,966 | 76.8 (70.6 to 82.1) | 12.8 (8.9 to 18.0) | 28.7 (22.8 to 35.5) | 4.4 (3.7 to 5.0) |
| Division | Chattogram | 5,145 | 78.5 (74.4 to 82.1) | 17.9 (14.3 to 22.2) | 32.0 (27.1 to 37.3) | 5.6 (4.9 to 6.4) |
| Division | Dhaka | 4,909 | 75.4 (71.6 to 78.9) | 37.0 (33.1 to 41.0) | 30.2 (26.6 to 34.1) | 13.3 (11.5 to 15.2) |
| Division | Khulna | 3,076 | 83.5 (79.1 to 87.1) | 17.5 (13.9 to 21.8) | 34.8 (29.8 to 40.1) | 5.8 (4.9 to 6.7) |
| Division | Mymensingh | 1,430 | 77.5 (69.6 to 83.9) | 14.1 (8.8 to 21.8) | 33.5 (25.6 to 42.5) | 4.5 (3.6 to 5.4) |
| Division | Rajshahi | 2,333 | 75.9 (70.5 to 80.5) | 16.3 (12.5 to 20.9) | 28.5 (23.6 to 33.9) | 5.2 (4.3 to 6.1) |
| Division | Rangpur | 2,843 | 72.2 (67.3 to 76.6) | 13.3 (9.9 to 17.7) | 31.5 (26.4 to 37.1) | 4.4 (3.8 to 5.0) |
| Division | Sylhet | 1,655 | 51.1 (43.4 to 58.8) | 3.4 (1.6 to 7.2) | 16.3 (11.7 to 22.3) | 1.9 (1.4 to 2.3) |
| Residence | Rural | 17,147 | 74.6 (72.5 to 76.6) | 16.4 (14.6 to 18.3) | 30.3 (28.0 to 32.7) | 5.9 (5.4 to 6.4) |
| Residence | Urban | 6,210 | 77.2 (73.7 to 80.3) | 29.9 (26.5 to 33.5) | 30.4 (27.0 to 34.1) | 9.4 (8.1 to 10.6) |
| Wealth quintile | Poorest | 5,441 | 67.8 (64.9 to 70.6) | 10.1 (8.5 to 12.1) | 27.1 (24.5 to 30.0) | 3.7 (3.4 to 4.1) |
| Wealth quintile | Second | 4,964 | 74.2 (71.7 to 76.5) | 12.3 (10.6 to 14.2) | 29.6 (27.0 to 32.4) | 4.4 (4.1 to 4.7) |
| Wealth quintile | Middle | 4,675 | 76.9 (74.5 to 79.2) | 19.4 (17.2 to 21.8) | 33.4 (30.7 to 36.2) | 6.7 (5.9 to 7.4) |
| Wealth quintile | Fourth | 4,410 | 77.9 (75.1 to 80.4) | 27.0 (24.2 to 29.9) | 32.3 (29.5 to 35.3) | 9.5 (8.4 to 10.7) |
| Wealth quintile | Richest | 3,867 | 80.0 (77.1 to 82.7) | 32.5 (29.3 to 35.8) | 29.1 (26.1 to 32.3) | 10.3 (9.1 to 11.4) |

Weighted estimates (95% CI) using child sample weights. Exposure is measured at the displaced cluster location with the main kiln map (Lee and colleagues, imagery from 2018 to 2019).

| Measure | Clusters | Value |
| --- | --- | --- |
| Share of the variation in the 5 km kiln count between clusters explained by district (R², %) | 3,145 | 26.7 |

From a linear regression of the cluster kiln count on district indicators.

### S3 Table. Adjusted prevalence ratios by period of interview or blood draw, and tests of interaction

| Outcome | Period | Category | Estimate | P for trend | n |
| --- | --- | --- | --- | --- | --- |
| Blood lead 5 µg/dL or more | April | 1 to 4 | 0.85 (0.73 to 1.00) | 0.921 | 2,931 |
| Blood lead 5 µg/dL or more | April | 5 to 9 | 1.06 (0.88 to 1.27) | 0.921 | 2,931 |
| Blood lead 5 µg/dL or more | April | 10 or more | 0.96 (0.82 to 1.13) | 0.921 | 2,931 |
| Blood lead 5 µg/dL or more | February to March | 1 to 4 | 0.88 (0.75 to 1.05) | 0.324 | 3,485 |
| Blood lead 5 µg/dL or more | February to March | 5 to 9 | 0.93 (0.77 to 1.13) | 0.324 | 3,485 |
| Blood lead 5 µg/dL or more | February to March | 10 or more | 0.87 (0.69 to 1.10) | 0.324 | 3,485 |
| Blood lead 5 µg/dL or more | May to June | 1 to 4 | 1.06 (0.93 to 1.21) | 0.599 | 4,202 |
| Blood lead 5 µg/dL or more | May to June | 5 to 9 | 1.07 (0.90 to 1.27) | 0.599 | 4,202 |
| Blood lead 5 µg/dL or more | May to June | 10 or more | 0.97 (0.83 to 1.14) | 0.599 | 4,202 |
| Cough | April | 1 to 4 | 1.07 (0.94 to 1.21) | 0.603 | 6,171 |
| Cough | April | 5 to 9 | 1.00 (0.85 to 1.17) | 0.603 | 6,171 |
| Cough | April | 10 or more | 0.93 (0.75 to 1.17) | 0.603 | 6,171 |
| Cough | February to March | 1 to 4 | 0.95 (0.85 to 1.05) | 0.627 | 8,075 |
| Cough | February to March | 5 to 9 | 0.98 (0.86 to 1.11) | 0.627 | 8,075 |
| Cough | February to March | 10 or more | 0.95 (0.83 to 1.09) | 0.627 | 8,075 |
| Cough | May to June | 1 to 4 | 1.07 (0.94 to 1.22) | 0.015 | 9,095 |
| Cough | May to June | 5 to 9 | 1.12 (0.96 to 1.31) | 0.015 | 9,095 |
| Cough | May to June | 10 or more | 1.24 (1.04 to 1.49) | 0.015 | 9,095 |
| Diarrhoea (negative control outcome) | April | 1 to 4 | 0.97 (0.77 to 1.23) | 0.941 | 6,172 |
| Diarrhoea (negative control outcome) | April | 5 to 9 | 1.08 (0.80 to 1.47) | 0.941 | 6,172 |
| Diarrhoea (negative control outcome) | April | 10 or more | 0.91 (0.61 to 1.34) | 0.941 | 6,172 |
| Diarrhoea (negative control outcome) | February to March | 1 to 4 | 1.04 (0.80 to 1.37) | 0.080 | 8,076 |
| Diarrhoea (negative control outcome) | February to March | 5 to 9 | 1.39 (1.05 to 1.85) | 0.080 | 8,076 |
| Diarrhoea (negative control outcome) | February to March | 10 or more | 1.23 (0.86 to 1.74) | 0.080 | 8,076 |
| Diarrhoea (negative control outcome) | May to June | 1 to 4 | 0.99 (0.73 to 1.35) | 0.888 | 9,091 |
| Diarrhoea (negative control outcome) | May to June | 5 to 9 | 1.13 (0.78 to 1.64) | 0.888 | 9,091 |
| Diarrhoea (negative control outcome) | May to June | 10 or more | 0.98 (0.67 to 1.42) | 0.888 | 9,091 |
| Symptoms of ARI | April | 1 to 4 | 1.25 (0.79 to 1.97) | 0.648 | 6,169 |
| Symptoms of ARI | April | 5 to 9 | 0.88 (0.49 to 1.60) | 0.648 | 6,169 |
| Symptoms of ARI | April | 10 or more | 0.90 (0.47 to 1.72) | 0.648 | 6,169 |
| Symptoms of ARI | February to March | 1 to 4 | 1.09 (0.77 to 1.55) | 0.592 | 8,073 |
| Symptoms of ARI | February to March | 5 to 9 | 0.95 (0.62 to 1.44) | 0.592 | 8,073 |
| Symptoms of ARI | February to March | 10 or more | 1.26 (0.79 to 2.02) | 0.592 | 8,073 |
| Symptoms of ARI | May to June | 1 to 4 | 0.90 (0.59 to 1.35) | 0.408 | 9,089 |
| Symptoms of ARI | May to June | 5 to 9 | 0.89 (0.54 to 1.47) | 0.408 | 9,089 |
| Symptoms of ARI | May to June | 10 or more | 1.22 (0.74 to 1.99) | 0.408 | 9,089 |

| Outcome | Test | F | P |
| --- | --- | --- | --- |
| Cough | Kiln category by period | 1.77 | 0.102 |
| Cough | Per 10 kilns by period | 3.92 | 0.020 |
| Symptoms of ARI | Kiln category by period | 0.47 | 0.830 |
| Symptoms of ARI | Per 10 kilns by period | 0.63 | 0.534 |
| Diarrhoea (negative control outcome) | Kiln category by period | 1.18 | 0.313 |
| Diarrhoea (negative control outcome) | Per 10 kilns by period | 1.38 | 0.253 |
| Blood lead 5 µg/dL or more | Kiln category by period | 2.25 | 0.036 |

Kilns fire in the dry season, from about November to April or May. February to March is the peak of firing; April is late in the season; May to June spans the end of firing (May) and the months after it (June). The February to March versus June comparison is in S1 Table. For blood outcomes the period is the month of the blood draw. Interaction tests: kiln category by period (6 df) and per 10 kilns by period (2 df).

### S4 Table. Stepwise adjustment

| Outcome | Model | Category | Estimate | n |
| --- | --- | --- | --- | --- |
| Cough | Child and household factors | 1 to 4 | 1.06 (0.98 to 1.14) | 23,341 |
| Cough | Child and household factors | 5 to 9 | 1.09 (1.00 to 1.19) | 23,341 |
| Cough | Child and household factors | 10 or more | 1.07 (0.98 to 1.17) | 23,341 |
| Cough | + urban, population density, night lights | 1 to 4 | 1.06 (0.98 to 1.14) | 23,341 |
| Cough | + urban, population density, night lights | 5 to 9 | 1.09 (1.00 to 1.19) | 23,341 |
| Cough | + urban, population density, night lights | 10 or more | 1.07 (0.98 to 1.18) | 23,341 |
| Cough | + division | 1 to 4 | 1.03 (0.96 to 1.10) | 23,341 |
| Cough | + division | 5 to 9 | 1.06 (0.98 to 1.16) | 23,341 |
| Cough | + division | 10 or more | 1.08 (0.99 to 1.18) | 23,341 |
| Cough | + district (main model) | 1 to 4 | 1.01 (0.94 to 1.08) | 23,341 |
| Cough | + district (main model) | 5 to 9 | 1.03 (0.95 to 1.12) | 23,341 |
| Cough | + district (main model) | 10 or more | 1.05 (0.96 to 1.16) | 23,341 |
| Symptoms of ARI | Child and household factors | 1 to 4 | 1.07 (0.85 to 1.34) | 23,331 |
| Symptoms of ARI | Child and household factors | 5 to 9 | 0.91 (0.69 to 1.20) | 23,331 |
| Symptoms of ARI | Child and household factors | 10 or more | 1.14 (0.87 to 1.50) | 23,331 |
| Symptoms of ARI | + urban, population density, night lights | 1 to 4 | 1.07 (0.85 to 1.35) | 23,331 |
| Symptoms of ARI | + urban, population density, night lights | 5 to 9 | 0.92 (0.70 to 1.21) | 23,331 |
| Symptoms of ARI | + urban, population density, night lights | 10 or more | 1.10 (0.84 to 1.44) | 23,331 |
| Symptoms of ARI | + division | 1 to 4 | 1.08 (0.86 to 1.36) | 23,331 |
| Symptoms of ARI | + division | 5 to 9 | 0.97 (0.73 to 1.28) | 23,331 |
| Symptoms of ARI | + division | 10 or more | 1.24 (0.95 to 1.62) | 23,331 |
| Symptoms of ARI | + district (main model) | 1 to 4 | 1.08 (0.86 to 1.36) | 23,331 |
| Symptoms of ARI | + district (main model) | 5 to 9 | 0.98 (0.73 to 1.33) | 23,331 |
| Symptoms of ARI | + district (main model) | 10 or more | 1.21 (0.90 to 1.62) | 23,331 |
| Blood lead 5 µg/dL or more | Child and household factors | 1 to 4 | 0.88 (0.81 to 0.97) | 10,618 |
| Blood lead 5 µg/dL or more | Child and household factors | 5 to 9 | 0.88 (0.79 to 0.99) | 10,618 |
| Blood lead 5 µg/dL or more | Child and household factors | 10 or more | 1.10 (1.00 to 1.21) | 10,618 |
| Blood lead 5 µg/dL or more | + urban, population density, night lights | 1 to 4 | 0.91 (0.83 to 1.00) | 10,618 |
| Blood lead 5 µg/dL or more | + urban, population density, night lights | 5 to 9 | 0.94 (0.84 to 1.05) | 10,618 |
| Blood lead 5 µg/dL or more | + urban, population density, night lights | 10 or more | 1.04 (0.95 to 1.14) | 10,618 |
| Blood lead 5 µg/dL or more | + division | 1 to 4 | 0.95 (0.87 to 1.03) | 10,618 |
| Blood lead 5 µg/dL or more | + division | 5 to 9 | 1.03 (0.93 to 1.14) | 10,618 |
| Blood lead 5 µg/dL or more | + division | 10 or more | 1.04 (0.96 to 1.14) | 10,618 |
| Blood lead 5 µg/dL or more | + district (main model) | 1 to 4 | 0.95 (0.87 to 1.03) | 10,618 |
| Blood lead 5 µg/dL or more | + district (main model) | 5 to 9 | 1.01 (0.91 to 1.12) | 10,618 |
| Blood lead 5 µg/dL or more | + district (main model) | 10 or more | 0.94 (0.85 to 1.03) | 10,618 |
| Log blood lead (difference) | Child and household factors | 1 to 4 | -0.04 (-0.08 to 0.00) | 10,618 |
| Log blood lead (difference) | Child and household factors | 5 to 9 | -0.05 (-0.09 to -0.00) | 10,618 |
| Log blood lead (difference) | Child and household factors | 10 or more | 0.07 (0.02 to 0.12) | 10,618 |
| Log blood lead (difference) | + urban, population density, night lights | 1 to 4 | -0.04 (-0.08 to 0.00) | 10,618 |
| Log blood lead (difference) | + urban, population density, night lights | 5 to 9 | -0.04 (-0.09 to 0.01) | 10,618 |
| Log blood lead (difference) | + urban, population density, night lights | 10 or more | 0.01 (-0.04 to 0.06) | 10,618 |
| Log blood lead (difference) | + division | 1 to 4 | -0.02 (-0.06 to 0.02) | 10,618 |
| Log blood lead (difference) | + division | 5 to 9 | 0.01 (-0.04 to 0.05) | 10,618 |
| Log blood lead (difference) | + division | 10 or more | 0.02 (-0.03 to 0.07) | 10,618 |
| Log blood lead (difference) | + district (main model) | 1 to 4 | -0.01 (-0.05 to 0.02) | 10,618 |
| Log blood lead (difference) | + district (main model) | 5 to 9 | 0.00 (-0.04 to 0.05) | 10,618 |
| Log blood lead (difference) | + district (main model) | 10 or more | -0.02 (-0.07 to 0.03) | 10,618 |

Child and household factors: age group, sex, mother’s education, wealth quintile, cooking fuel and month. Crude estimates are in S5 Table.

### S5 Table. Crude and adjusted associations of kiln category with all outcomes

| Outcome | Model | Category | Estimate | P | P for trend | n |
| --- | --- | --- | --- | --- | --- | --- |
| Cough | Crude | 1 to 4 | 1.08 (1.00 to 1.17) | 0.056 | 0.090 | 23,341 |
| Cough | Crude | 5 to 9 | 1.10 (1.01 to 1.20) | 0.034 | 0.090 | 23,341 |
| Cough | Crude | 10 or more | 1.08 (0.98 to 1.18) | 0.104 | 0.090 | 23,341 |
| Cough | Adjusted | 1 to 4 | 1.01 (0.94 to 1.08) | 0.741 | 0.242 | 23,341 |
| Cough | Adjusted | 5 to 9 | 1.03 (0.95 to 1.12) | 0.429 | 0.242 | 23,341 |
| Cough | Adjusted | 10 or more | 1.05 (0.96 to 1.16) | 0.280 | 0.242 | 23,341 |
| Symptoms of ARI | Crude | 1 to 4 | 1.10 (0.87 to 1.38) | 0.426 | 0.766 | 23,331 |
| Symptoms of ARI | Crude | 5 to 9 | 0.93 (0.71 to 1.23) | 0.625 | 0.766 | 23,331 |
| Symptoms of ARI | Crude | 10 or more | 1.11 (0.84 to 1.45) | 0.463 | 0.766 | 23,331 |
| Symptoms of ARI | Adjusted | 1 to 4 | 1.08 (0.86 to 1.36) | 0.510 | 0.374 | 23,331 |
| Symptoms of ARI | Adjusted | 5 to 9 | 0.98 (0.73 to 1.33) | 0.914 | 0.374 | 23,331 |
| Symptoms of ARI | Adjusted | 10 or more | 1.21 (0.90 to 1.62) | 0.208 | 0.374 | 23,331 |
| Diarrhoea (negative control outcome) | Crude | 1 to 4 | 1.01 (0.86 to 1.19) | 0.895 | 0.751 | 23,339 |
| Diarrhoea (negative control outcome) | Crude | 5 to 9 | 1.18 (0.99 to 1.41) | 0.061 | 0.751 | 23,339 |
| Diarrhoea (negative control outcome) | Crude | 10 or more | 0.97 (0.81 to 1.16) | 0.711 | 0.751 | 23,339 |
| Diarrhoea (negative control outcome) | Adjusted | 1 to 4 | 0.96 (0.82 to 1.13) | 0.654 | 0.565 | 23,339 |
| Diarrhoea (negative control outcome) | Adjusted | 5 to 9 | 1.17 (0.97 to 1.40) | 0.099 | 0.565 | 23,339 |
| Diarrhoea (negative control outcome) | Adjusted | 10 or more | 0.97 (0.80 to 1.19) | 0.798 | 0.565 | 23,339 |
| Blood lead 5 µg/dL or more | Crude | 1 to 4 | 0.84 (0.76 to 0.94) | 0.002 | <0.001 | 10,618 |
| Blood lead 5 µg/dL or more | Crude | 5 to 9 | 0.83 (0.73 to 0.93) | 0.002 | <0.001 | 10,618 |
| Blood lead 5 µg/dL or more | Crude | 10 or more | 1.27 (1.14 to 1.41) | <0.001 | <0.001 | 10,618 |
| Blood lead 5 µg/dL or more | Adjusted | 1 to 4 | 0.95 (0.87 to 1.03) | 0.192 | 0.403 | 10,618 |
| Blood lead 5 µg/dL or more | Adjusted | 5 to 9 | 1.01 (0.91 to 1.12) | 0.873 | 0.403 | 10,618 |
| Blood lead 5 µg/dL or more | Adjusted | 10 or more | 0.94 (0.85 to 1.03) | 0.194 | 0.403 | 10,618 |
| Blood lead 5 µg/dL or more | Adjusted plus lead sources | 1 to 4 | 0.94 (0.87 to 1.03) | 0.176 | 0.391 | 10,612 |
| Blood lead 5 µg/dL or more | Adjusted plus lead sources | 5 to 9 | 1.01 (0.91 to 1.12) | 0.868 | 0.391 | 10,612 |
| Blood lead 5 µg/dL or more | Adjusted plus lead sources | 10 or more | 0.93 (0.84 to 1.03) | 0.182 | 0.391 | 10,612 |
| Blood lead 3.5 µg/dL or more | Crude | 1 to 4 | 0.94 (0.89 to 1.00) | 0.038 | 0.084 | 10,618 |
| Blood lead 3.5 µg/dL or more | Crude | 5 to 9 | 0.89 (0.83 to 0.95) | <0.001 | 0.084 | 10,618 |
| Blood lead 3.5 µg/dL or more | Crude | 10 or more | 1.08 (1.02 to 1.15) | 0.006 | 0.084 | 10,618 |
| Blood lead 3.5 µg/dL or more | Adjusted | 1 to 4 | 1.00 (0.95 to 1.04) | 0.934 | 0.146 | 10,618 |
| Blood lead 3.5 µg/dL or more | Adjusted | 5 to 9 | 0.98 (0.93 to 1.04) | 0.559 | 0.146 | 10,618 |
| Blood lead 3.5 µg/dL or more | Adjusted | 10 or more | 0.96 (0.91 to 1.02) | 0.159 | 0.146 | 10,618 |
| Blood lead 3.5 µg/dL or more | Adjusted plus lead sources | 1 to 4 | 1.00 (0.95 to 1.04) | 0.920 | 0.145 | 10,612 |
| Blood lead 3.5 µg/dL or more | Adjusted plus lead sources | 5 to 9 | 0.98 (0.93 to 1.04) | 0.568 | 0.145 | 10,612 |
| Blood lead 3.5 µg/dL or more | Adjusted plus lead sources | 10 or more | 0.96 (0.91 to 1.02) | 0.154 | 0.145 | 10,612 |
| Log blood lead (difference) | Crude | 1 to 4 | -0.07 (-0.11 to -0.02) | 0.004 | <0.001 | 10,618 |
| Log blood lead (difference) | Crude | 5 to 9 | -0.08 (-0.13 to -0.03) | 0.001 | <0.001 | 10,618 |
| Log blood lead (difference) | Crude | 10 or more | 0.14 (0.08 to 0.20) | <0.001 | <0.001 | 10,618 |
| Log blood lead (difference) | Adjusted | 1 to 4 | -0.01 (-0.05 to 0.02) | 0.462 | 0.637 | 10,618 |
| Log blood lead (difference) | Adjusted | 5 to 9 | 0.00 (-0.04 to 0.05) | 0.843 | 0.637 | 10,618 |
| Log blood lead (difference) | Adjusted | 10 or more | -0.02 (-0.07 to 0.03) | 0.425 | 0.637 | 10,618 |
| Log blood lead (difference) | Adjusted plus lead sources | 1 to 4 | -0.01 (-0.05 to 0.02) | 0.466 | 0.625 | 10,612 |
| Log blood lead (difference) | Adjusted plus lead sources | 5 to 9 | 0.00 (-0.04 to 0.05) | 0.828 | 0.625 | 10,612 |
| Log blood lead (difference) | Adjusted plus lead sources | 10 or more | -0.02 (-0.07 to 0.03) | 0.410 | 0.625 | 10,612 |

Estimates are prevalence ratios for binary outcomes and differences for log blood lead (exponentiate for the ratio of geometric means). Reference: no kiln within 5 km. Lead sources: smoking by a person in regular contact with the child, pica, lead-related activity near the home, a household contact working with lead, and cosmetic use (sindoor or surma).

### S6 Table. Household soil lead and exploratory blood metals by kiln category

| Measure | Households | Value |
| --- | --- | --- |
| Household soil lead, weighted geometric mean (mg/kg) | 10,550 | 21.5 |

| Outcome | Model | Category | Difference (95% CI) | Ratio of geometric means | P for trend | n |
| --- | --- | --- | --- | --- | --- | --- |
| Log household soil lead | Crude | 1 to 4 | 0.02 (-0.02 to 0.06) | 1.02 (0.98 to 1.07) | 0.001 | 10,546 |
| Log household soil lead | Crude | 5 to 9 | -0.01 (-0.06 to 0.03) | 0.99 (0.94 to 1.03) | 0.001 | 10,546 |
| Log household soil lead | Crude | 10 or more | 0.11 (0.05 to 0.16) | 1.11 (1.06 to 1.17) | 0.001 | 10,546 |
| Log household soil lead | Adjusted | 1 to 4 | 0.03 (-0.01 to 0.06) | 1.03 (0.99 to 1.06) | 0.629 | 10,546 |
| Log household soil lead | Adjusted | 5 to 9 | 0.01 (-0.03 to 0.04) | 1.01 (0.97 to 1.04) | 0.629 | 10,546 |
| Log household soil lead | Adjusted | 10 or more | -0.01 (-0.05 to 0.04) | 0.99 (0.95 to 1.04) | 0.629 | 10,546 |
| Log blood cadmium (exploratory) | Adjusted | 1 to 4 | -0.08 (-0.13 to -0.03) | 0.92 (0.88 to 0.97) | <0.001 | 10,619 |
| Log blood cadmium (exploratory) | Adjusted | 5 to 9 | -0.08 (-0.15 to -0.01) | 0.92 (0.86 to 0.99) | <0.001 | 10,619 |
| Log blood cadmium (exploratory) | Adjusted | 10 or more | -0.18 (-0.26 to -0.10) | 0.84 (0.77 to 0.91) | <0.001 | 10,619 |
| Log blood arsenic (exploratory) | Adjusted | 1 to 4 | 0.00 (-0.06 to 0.06) | 1.00 (0.95 to 1.06) | 0.759 | 10,620 |
| Log blood arsenic (exploratory) | Adjusted | 5 to 9 | 0.01 (-0.06 to 0.08) | 1.01 (0.94 to 1.09) | 0.759 | 10,620 |
| Log blood arsenic (exploratory) | Adjusted | 10 or more | 0.01 (-0.07 to 0.09) | 1.01 (0.93 to 1.09) | 0.759 | 10,620 |
| Log blood mercury (exploratory) | Adjusted | 1 to 4 | -0.03 (-0.08 to 0.02) | 0.97 (0.92 to 1.02) | 0.459 | 10,620 |
| Log blood mercury (exploratory) | Adjusted | 5 to 9 | -0.02 (-0.08 to 0.04) | 0.98 (0.92 to 1.04) | 0.459 | 10,620 |
| Log blood mercury (exploratory) | Adjusted | 10 or more | -0.03 (-0.10 to 0.04) | 0.97 (0.90 to 1.04) | 0.459 | 10,620 |

The survey report did not publish cadmium, arsenic and mercury, because detection thresholds, analytical precision and reference thresholds did not allow robust estimates, so these results are exploratory. Soil was sampled only in blood-module households, so soil lead and blood metals use blood sample weights and the blood adjustment set (month of blood draw).

### S7 Table. Sensitivity analyses: other exposure measures, a second kiln map and urban clusters

| Analysis | Map | Outcome | Level | Estimate | P for trend | n |
| --- | --- | --- | --- | --- | --- | --- |
| Nearest kiln distance (reference 10 km or more) | Main map | Cough | 5 to 10 km | 1.30 (1.13 to 1.50) | 0.167 | 23,341 |
| Nearest kiln distance (reference 10 km or more) | Main map | Cough | 2 to 5 km | 1.28 (1.12 to 1.48) | 0.167 | 23,341 |
| Nearest kiln distance (reference 10 km or more) | Main map | Cough | Under 2 km | 1.28 (1.11 to 1.48) | 0.167 | 23,341 |
| Kilns within 10 km (reference none) | Main map | Cough | 1 to 9 | 1.26 (1.10 to 1.44) | 0.004 | 23,341 |
| Kilns within 10 km (reference none) | Main map | Cough | 10 to 29 | 1.33 (1.15 to 1.53) | 0.004 | 23,341 |
| Kilns within 10 km (reference none) | Main map | Cough | 30 or more | 1.35 (1.15 to 1.57) | 0.004 | 23,341 |
| Nearest kiln distance (reference 10 km or more) | Main map | Symptoms of ARI | 5 to 10 km | 1.16 (0.73 to 1.82) | 0.474 | 23,331 |
| Nearest kiln distance (reference 10 km or more) | Main map | Symptoms of ARI | 2 to 5 km | 1.22 (0.78 to 1.90) | 0.474 | 23,331 |
| Nearest kiln distance (reference 10 km or more) | Main map | Symptoms of ARI | Under 2 km | 1.22 (0.77 to 1.93) | 0.474 | 23,331 |
| Kilns within 10 km (reference none) | Main map | Symptoms of ARI | 1 to 9 | 1.14 (0.74 to 1.75) | 0.290 | 23,331 |
| Kilns within 10 km (reference none) | Main map | Symptoms of ARI | 10 to 29 | 1.31 (0.82 to 2.11) | 0.290 | 23,331 |
| Kilns within 10 km (reference none) | Main map | Symptoms of ARI | 30 or more | 1.27 (0.76 to 2.11) | 0.290 | 23,331 |
| Nearest kiln distance (reference 10 km or more) | Main map | Blood lead 5 µg/dL or more | 5 to 10 km | 1.03 (0.88 to 1.22) | 0.607 | 10,618 |
| Nearest kiln distance (reference 10 km or more) | Main map | Blood lead 5 µg/dL or more | 2 to 5 km | 0.98 (0.83 to 1.14) | 0.607 | 10,618 |
| Nearest kiln distance (reference 10 km or more) | Main map | Blood lead 5 µg/dL or more | Under 2 km | 1.00 (0.85 to 1.17) | 0.607 | 10,618 |
| Kilns within 10 km (reference none) | Main map | Blood lead 5 µg/dL or more | 1 to 9 | 0.98 (0.84 to 1.15) | 0.551 | 10,618 |
| Kilns within 10 km (reference none) | Main map | Blood lead 5 µg/dL or more | 10 to 29 | 1.03 (0.87 to 1.21) | 0.551 | 10,618 |
| Kilns within 10 km (reference none) | Main map | Blood lead 5 µg/dL or more | 30 or more | 1.02 (0.85 to 1.23) | 0.551 | 10,618 |
| Six divisions mapped by both sources | Second map (APAD) | Cough | 1 to 4 | 0.99 (0.90 to 1.09) | 0.454 | 16,545 |
| Six divisions mapped by both sources | Second map (APAD) | Cough | 5 to 9 | 0.92 (0.82 to 1.03) | 0.454 | 16,545 |
| Six divisions mapped by both sources | Second map (APAD) | Cough | 10 or more | 0.98 (0.86 to 1.12) | 0.454 | 16,545 |
| Six divisions mapped by both sources | Main map | Cough | 1 to 4 | 1.01 (0.92 to 1.11) | 0.651 | 16,545 |
| Six divisions mapped by both sources | Main map | Cough | 5 to 9 | 1.06 (0.95 to 1.17) | 0.651 | 16,545 |
| Six divisions mapped by both sources | Main map | Cough | 10 or more | 1.01 (0.90 to 1.14) | 0.651 | 16,545 |
| Six divisions mapped by both sources | Second map (APAD) | Symptoms of ARI | 1 to 4 | 1.01 (0.69 to 1.48) | 0.550 | 16,546 |
| Six divisions mapped by both sources | Second map (APAD) | Symptoms of ARI | 5 to 9 | 0.79 (0.51 to 1.22) | 0.550 | 16,546 |
| Six divisions mapped by both sources | Second map (APAD) | Symptoms of ARI | 10 or more | 0.97 (0.61 to 1.55) | 0.550 | 16,546 |
| Six divisions mapped by both sources | Main map | Symptoms of ARI | 1 to 4 | 0.94 (0.69 to 1.29) | 0.547 | 16,546 |
| Six divisions mapped by both sources | Main map | Symptoms of ARI | 5 to 9 | 0.75 (0.50 to 1.12) | 0.547 | 16,546 |
| Six divisions mapped by both sources | Main map | Symptoms of ARI | 10 or more | 0.96 (0.66 to 1.40) | 0.547 | 16,546 |
| Six divisions mapped by both sources | Second map (APAD) | Blood lead 5 µg/dL or more | 1 to 4 | 0.96 (0.86 to 1.08) | 0.287 | 7,820 |
| Six divisions mapped by both sources | Second map (APAD) | Blood lead 5 µg/dL or more | 5 to 9 | 0.99 (0.87 to 1.14) | 0.287 | 7,820 |
| Six divisions mapped by both sources | Second map (APAD) | Blood lead 5 µg/dL or more | 10 or more | 0.91 (0.78 to 1.05) | 0.287 | 7,820 |
| Six divisions mapped by both sources | Main map | Blood lead 5 µg/dL or more | 1 to 4 | 0.97 (0.87 to 1.08) | 0.601 | 7,820 |
| Six divisions mapped by both sources | Main map | Blood lead 5 µg/dL or more | 5 to 9 | 1.05 (0.92 to 1.19) | 0.601 | 7,820 |
| Six divisions mapped by both sources | Main map | Blood lead 5 µg/dL or more | 10 or more | 1.01 (0.90 to 1.13) | 0.601 | 7,820 |
| Urban clusters only | Main map | Cough | 1 to 4 | 1.05 (0.92 to 1.20) | 0.407 | 6,208 |
| Urban clusters only | Main map | Cough | 5 to 9 | 1.13 (0.95 to 1.34) | 0.407 | 6,208 |
| Urban clusters only | Main map | Cough | 10 or more | 1.07 (0.90 to 1.27) | 0.407 | 6,208 |
| Urban clusters only | Main map | Symptoms of ARI | 1 to 4 | 1.56 (0.96 to 2.52) | 0.007 | 6,201 |
| Urban clusters only | Main map | Symptoms of ARI | 5 to 9 | 1.27 (0.67 to 2.41) | 0.007 | 6,201 |
| Urban clusters only | Main map | Symptoms of ARI | 10 or more | 2.37 (1.33 to 4.21) | 0.007 | 6,201 |
| Urban clusters only | Main map | Blood lead 5 µg/dL or more | 1 to 4 | 0.92 (0.80 to 1.06) | 0.783 | 2,639 |
| Urban clusters only | Main map | Blood lead 5 µg/dL or more | 5 to 9 | 1.16 (0.98 to 1.38) | 0.783 | 2,639 |
| Urban clusters only | Main map | Blood lead 5 µg/dL or more | 10 or more | 0.98 (0.87 to 1.11) | 0.783 | 2,639 |

All models use the main adjustment set.

| Division | Children with no kiln within 10 km | % of this group |
| --- | --- | --- |
| Chattogram | 471 | 29.8 |
| Sylhet | 313 | 19.8 |
| Rangpur | 294 | 18.6 |
| Rajshahi | 157 | 9.9 |
| Dhaka | 107 | 6.8 |
| Mymensingh | 90 | 5.7 |
| Barishal | 82 | 5.2 |
| Khulna | 64 | 4.1 |

Unweighted counts by division of the children with no kiln within 10 km (the reference group for the 10 km count), who are 5.2% of all children (weighted).

| Kilns within 5 km | Urban children analysed | Children with symptoms of ARI |
| --- | --- | --- |
| None | 1,438 | 34 |
| 1 to 4 | 2,115 | 60 |
| 5 to 9 | 1,041 | 32 |
| 10 or more | 1,607 | 49 |

Unweighted counts in the urban-only analysis.

### S8 Table. Stability of kiln exposure under a second random displacement, and agreement between the two kiln maps

| Measure | Mean over 100 runs | 2.5th to 97.5th percentile of runs |
| --- | --- | --- |
| Spearman correlation of 5 km kiln count, all clusters | 0.86 | 0.85 to 0.87 |
| Spearman correlation, urban clusters | 0.94 | 0.92 to 0.95 |
| Spearman correlation, rural clusters | 0.82 | 0.81 to 0.84 |
| Same kiln category, all clusters (proportion) | 0.75 | 0.74 to 0.77 |
| Same kiln category, urban clusters | 0.85 | 0.83 to 0.87 |
| Same kiln category, rural clusters | 0.71 | 0.70 to 0.73 |
| Clusters with 10 or more kilns that stay in that category | 0.81 | 0.78 to 0.83 |
| Clusters with no kiln that stay with none | 0.83 | 0.81 to 0.85 |
| Agreement on nearest kiln under 2 km | 0.81 | 0.80 to 0.82 |
| Slope of re-displaced on published 5 km count | 0.89 | 0.81 to 0.95 |
| Pearson correlation of 5 km count within districts (district means removed) | 0.85 | 0.80 to 0.89 |
| Slope of re-displaced on published 5 km count within districts | 0.86 | 0.78 to 0.92 |

| Comparison | Kilns in Bangladesh | Matched within 500 m | % |
| --- | --- | --- | --- |
| Lee 2021 kilns matched by APAD | 6,976 | 3,258 | 46.7 |
| APAD kilns matched by Lee 2021 | 4,758 | 3,085 | 64.8 |

| Measure | Clusters | Value |
| --- | --- | --- |
| Spearman correlation of 5 km kiln counts, main versus second map (clusters in the six divisions covered by both) | 2,393 | 0.56 |

Each published cluster point was displaced again with the survey rule (urban up to 2 km; rural up to 5 km, 1% up to 10 km), 100 times, and exposure was recomputed. Percentiles describe the spread across runs and are not confidence intervals. No upazila or land constraint was applied. The APAD map does not cover Chattogram and Sylhet divisions.

### S1 Fig. Number of kilns within 5 km and child outcomes, restricted cubic spline

Ratio (95% CI) of each outcome at 1 to 55 kilns within 5 km compared with no kiln, from survey-weighted models with the main adjustment set. The count is modelled with an indicator for any kiln and a restricted cubic spline among clusters with at least one kiln (knots at 1, 4 and 16 kilns, the 10th, 50th and 90th centiles of non-zero counts). Prevalence ratios for binary outcomes; ratio of geometric means for blood lead. P values test any association (3 df) and non-linearity among exposed clusters (1 df). The curve stops at 55 kilns, the 99th centile; 60 of 3,149 clusters had 42 or more. Added after review of a first draft.

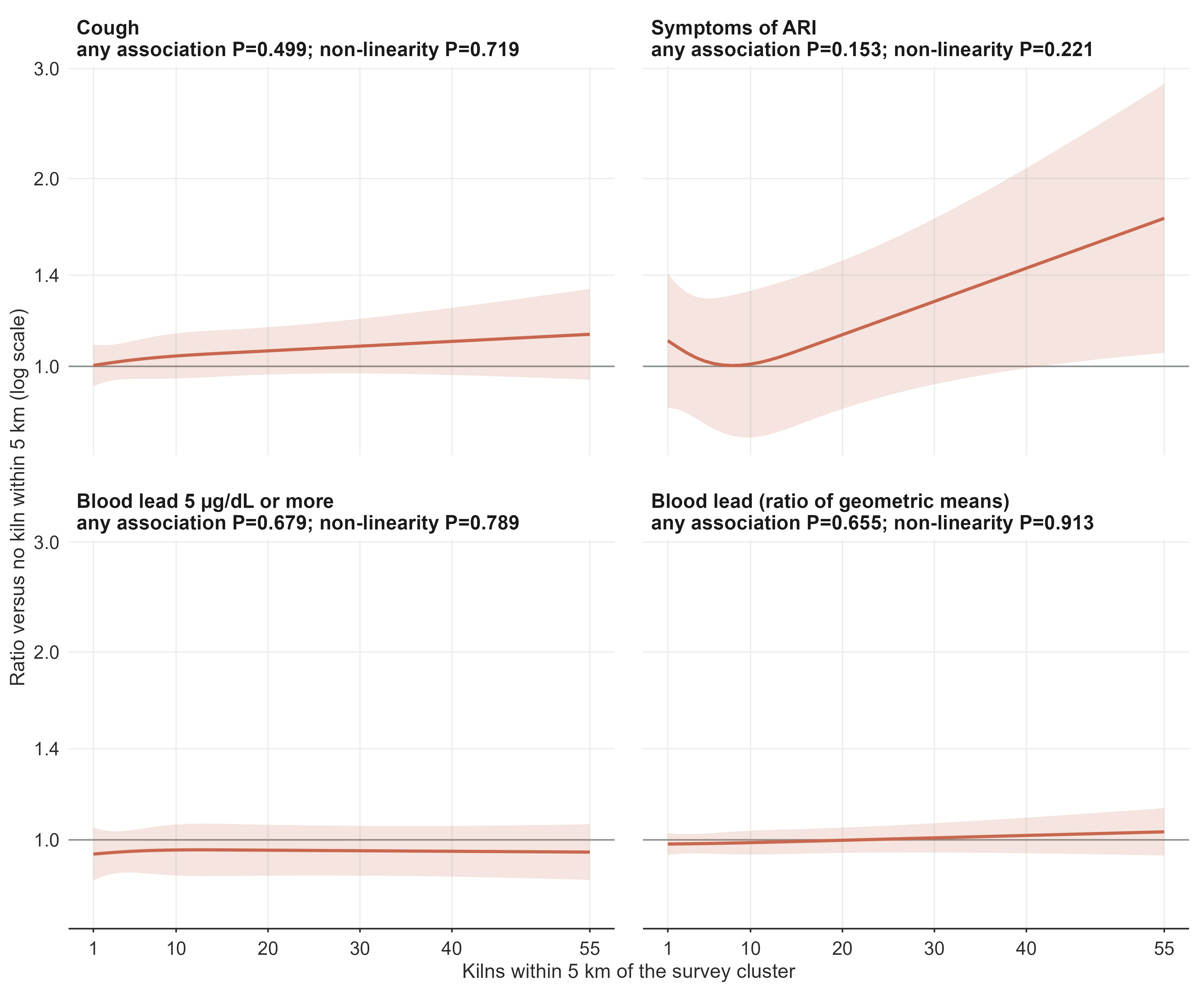

### S2 Fig. Stepwise adjustment of the association between ten or more kilns within 5 km and each outcome

Prevalence ratios with 95% CIs as covariates are added in steps. The final row is the main model.

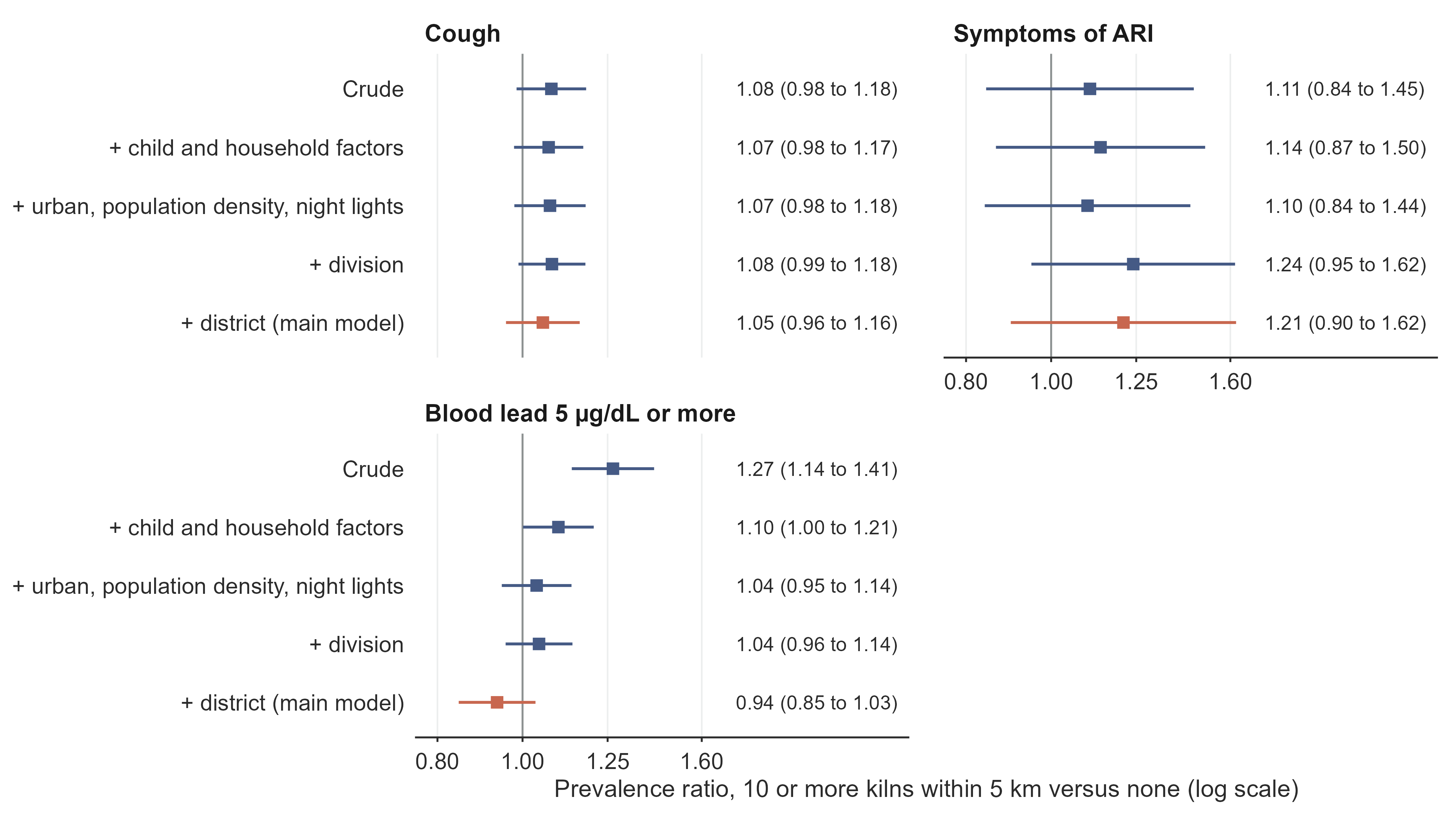
